# Exploring Machine Learning Models to Uncover Pathways in ALS Pathogenesis Using Immunohistochemical Features

**DOI:** 10.64898/2025.12.19.25342356

**Authors:** Jemimah Maria Kuruvilla, Olivia M Rifai, James Longden, Jenna M Gregory, Marta Vallejo

## Abstract

Hexanucleotide repeat expansions in *C9orf72* are the most common genetic cause of amyotrophic lateral sclerosis (ALS) and frontotemporal dementia, yet the predictive information contained within associated inflammatory and proteinopathy markers remains incompletely characterised. Here, we reconstructed five machine-learning-ready datasets from quantitative post-mortem measurements of Iba1, CD68, GFAP, TDP-43 and FUS obtained from 10 C9orf72-ALS cases and 10 controls, building upon the digital pathology study of Rifai *et al.*. Random forest, support vector machine, XGBoost, logistic regression, artificial neural network and ensemble approaches were evaluated, together with preprocessing, hyperparameter optimisation and biomarker-specific feature importance. Under conventional 3-fold cross-validation, model performance varied according to both biomarker and classifier, with no single algorithm consistently achieving the highest sensitivity, specificity and accuracy across all biomarkers. Iba1 showed the strongest random forest performance, reaching 88% sensitivity and 83% specificity. However, when random forest was evaluated using patient-grouped 5-fold cross-validation, performance decreased across all five biomarkers, with reductions of 6.98-14.52% in accuracy, 5.68-18.90% in sensitivity and 2.86-13.04% in specificity relative to conventional cross-validation. Iba1 nevertheless remained the strongest-performing random forest biomarker under patient-grouped evaluation. Feature-importance analysis further showed biomarker-specific predictive profiles, with classification generally distributed across multiple pathological measurements rather than dominated by a single feature. These findings demonstrate that estimates of machine-learning performance in quantitative neuropathology are sensitive to validation design and highlight the importance of patient-level separation when multiple pathological observations originate from the same individuals. Further evaluation using fully nested patient-grouped cross-validation is required to establish comparative model performance on unseen patients.

## 1 Introduction

Amyotrophic Lateral Sclerosis (ALS) is a motor neuron disease that causes progressive muscle wasting and paralysis, leading to loss of muscle endurance^1,2^. It predominantly degenerates motor neurons in the spinal, bulbar, and cortical regions^3^. Regardless of age, it is always terminal, with a life expectancy of 2-5 years^4^. The fatality of the disease is linked to certain factors like age, region of the brain affected, and genetic disposition, among others^5^. Early detection of ALS is essential for providing individualised care, optimising medical treatment, and potentially extending life expectancy^6^. ALS patients experience significant physical and psychological disability due to paralysis and loss of independence to perform daily tasks, placing a significant burden on both the patient and their caregivers^7^. According to a systematic review of cost-of-illness studies, ALS imposes a substantial economic burden on both patients and healthcare systems, with costs increasing as the disease progresses and peaking during the final stages of life^8^.

Familial ALS (fALS) is identified clinically by a family history consistent with inherited genetic patterns, most commonly autosomal dominant, with autosomal recessive and X-linked inheritance reported only rarely^9,10^. Sporadic ALS (sALS), by contrast, occurs without a known family history. However, a proportion of patients with sALS carry pathogenic or risk-associated variants in the same genes implicated in fALS. This overlap may reflect incomplete family records, small family sizes, or reduced gene penetrance. As genetic testing becomes more widely adopted, the traditional distinction between fALS and sALS may increasingly shift towards a classification based on genetic findings rather than family history alone^9,10^.

The genetic and molecular mechanisms underlying ALS remain incompletely understood. Several ALS-associated genes and pathological proteins, including TAR DNA-binding protein 43 (TDP-43) and Fused in Sarcoma (FUS), have been implicated in disease pathogenesis and contribute to the marked clinical and pathological heterogeneity of ALS. The hexanucleotide repeat expansion (HRE) in the *C9orf72* gene is the most common monogenic cause shared by both familial and sporadic ALS^9^. Although the precise biological function of C9orf72 remains incompletely understood, it has been implicated in intracellular protein homeostasis and vesicle trafficking^11^. Expansion of the repeat is associated with a spectrum of neurodegenerative phenotypes, including ALS-frontotemporal spectrum disorders (ALS-FTSD)^12^. ALS-FTSD encompasses a continuum of cognitive and behavioural impairment ranging from subtle deficits to clinically overt frontotemporal dementia (FTD), highlighting that ALS extends beyond motor neuron degeneration^11^. This clinical and pathological heterogeneity complicates therapeutic development, as clinical trials frequently rely on phenotypic outcome measures, including survival and motor and cognitive function, which vary considerably between patients^13^. Current diagnosis relies on clinical assessment supported by electrophysiology, neuroimaging, laboratory investigations, genetic testing and biomarker evaluation^14^. Despite these advances, objective pathological biomarkers capable of improving patient stratification and providing deeper insight into disease mechanisms remain limited.

Recent developments in machine learning (ML) have created new opportunities to improve disease identification and management by enabling the analysis of complex, high-dimensional data and revealing patterns that conventional analytical approaches may overlook. In ALS, ML has been applied to predict disease onset, progression and clinical outcomes, as well as to identify genetic and molecular factors associated with disease heterogeneity^15^. These advances have been supported by large publicly available resources, including the Pooled Resource Open-Access ALS Clinical Trials Database (PRO-ACT)^16^, the Answer ALS initiative^17^ and Project MinE^18^, which have substantially advanced clinical, molecular and genetic studies of ALS. However, these resources primarily comprise clinical, genetic or multi-omics data, whereas publicly available datasets containing quantitative post-mortem neuropathological measurements linked to detailed clinicopathological information remain comparatively scarce^19^. This limits the rigorous validation of computational approaches for tissue-level biomarker discovery and restricts our understanding of how pathological changes relate to clinical phenotypes^11^. Digital pathology addresses this gap by enabling the quantitative analysis of post-mortem tissue to characterise cellular alterations and proteinopathies associated with neurodegeneration^11,20^. Building on the work of Rifai *et al.*^11^, this study analyses the same post-mortem C9orf72-ALS dataset using an expanded and rigorously validated machine learning framework to identify reproducible pathological biomarkers and phenotypic signatures associated with C9orf72-associated ALS.

Quantitative immunohistochemical markers of neuroinflammation and proteinopathy are used to identify reproducible pathological signatures associated with disease status and biomarker prioritisation. Specifically, we aim to determine which protein- and cell-level markers most effectively distinguish ALS from control samples. Through the systematic evaluation of pathological patterns, this approach provides insights into the molecular and cellular processes underlying neurodegeneration. This study is not designed as a diagnostic or early-detection tool, as it relies on postmortem samples rather than data from living patients. In contrast, early detection of neurodegeneration has been demonstrated in a study by Langerova *et al.*^20^ on gastrointestinal pathology, which showed that early gut protein misfolding occurred years before ALS diagnosis. The present study differs by focusing on post-mortem brain tissue to understand disease mechanisms and evaluate ML frameworks, rather than predicting disease onset.

Building upon the work of Rifai et al.^11^, this study establishes a reproducible analytical framework for the computational analysis of quantitative neuropathology in C9orf72-associated ALS. The proposed framework integrates systematic prepro- cessing assessment, biomarker-specific hyperparameter optimisation, patient-level validation to eliminate data leakage, and feature importance analysis to improve the robustness, reproducibility and biological interpretability of machine learning models. The analytical pipeline is benchmarked directly against the original study using a broader range of supervised learning algorithms and consistent evaluation metrics. In addition, curated tabular datasets are generated for each biomarker to facilitate reproducible analyses and future methodological development. Rather than focusing solely on predictive performance, this work aims to provide a more rigorous and interpretable evaluation of disease-associated pathological signatures, establishing a computational framework that supports biomarker prioritisation and future computational pathology studies in ALS.

## 2 Results

In this section, results are presented following the pre-processing and ML pipelines outlined in the appendix, as shown in Figures 2 and 3, respectively.

### Data Pre-processing Results

To evaluate whether zero-variance features influenced model performance, models were trained with and without these features. Their removal did not produce consistent improvements across biomarkers and, in several cases, resulted in reduced classification performance (Tables 1 and 13). For example, Iba1 classification accuracy decreased by 1.17%, while TDP43 specificity decreased by 11.29% following feature removal. Given the absence of systematic performance gains and to maintain direct comparability with Rifai *et al.*^11^, zero-variance features were retained in the final analytical pipeline.

**Table 1.** Evaluation of feature-reduction strategies. Percentage change in the random forest classification performance after removing constant and collinear features relative to the complete feature set. Positive values indicate improved performance, whereas negative values indicate reduced performance.

| Biomarker | Constant features removed |  |  | Collinear features removed |  |  |
| --- | --- | --- | --- | --- | --- | --- |
|  | Accuracy (%) | Sensitivity (%) | Specificity (%) | Accuracy (%) | Sensitivity (%) | Specificity (%) |
| FUS | 0.00 | -1.39 | +1.33 | -2.82 | -1.37 | -4.23 |
| CD68 | 0.00 | +3.22 | -2.77 | +1.42 | +4.61 | -2.70 |
| Iba1 | -1.17 | 0.00 | 0.00 | 0.00 | -4.67 | +4.81 |
| TDP43 | -4.76 | +4.61 | -11.29 | -3.12 | -6.89 | +1.42 |
| GFAP | 0.00 | +2.77 | -4.47 | 0.00 | -1.44 | +2.77 |

Correlation analysis identified highly correlated features across all biomarker datasets (Figures 4–8). However, removing these features did not produce consistent improvements in classification performance (Table 1). Although modest gains were observed for individual performance metrics in some biomarkers, these were offset by reductions in others, resulting in no systematic improvement in overall model performance. Consequently, highly correlated features were retained in the final analytical pipeline. The complete random forest benchmarking results are presented in Table 16.

**Figure 1.**
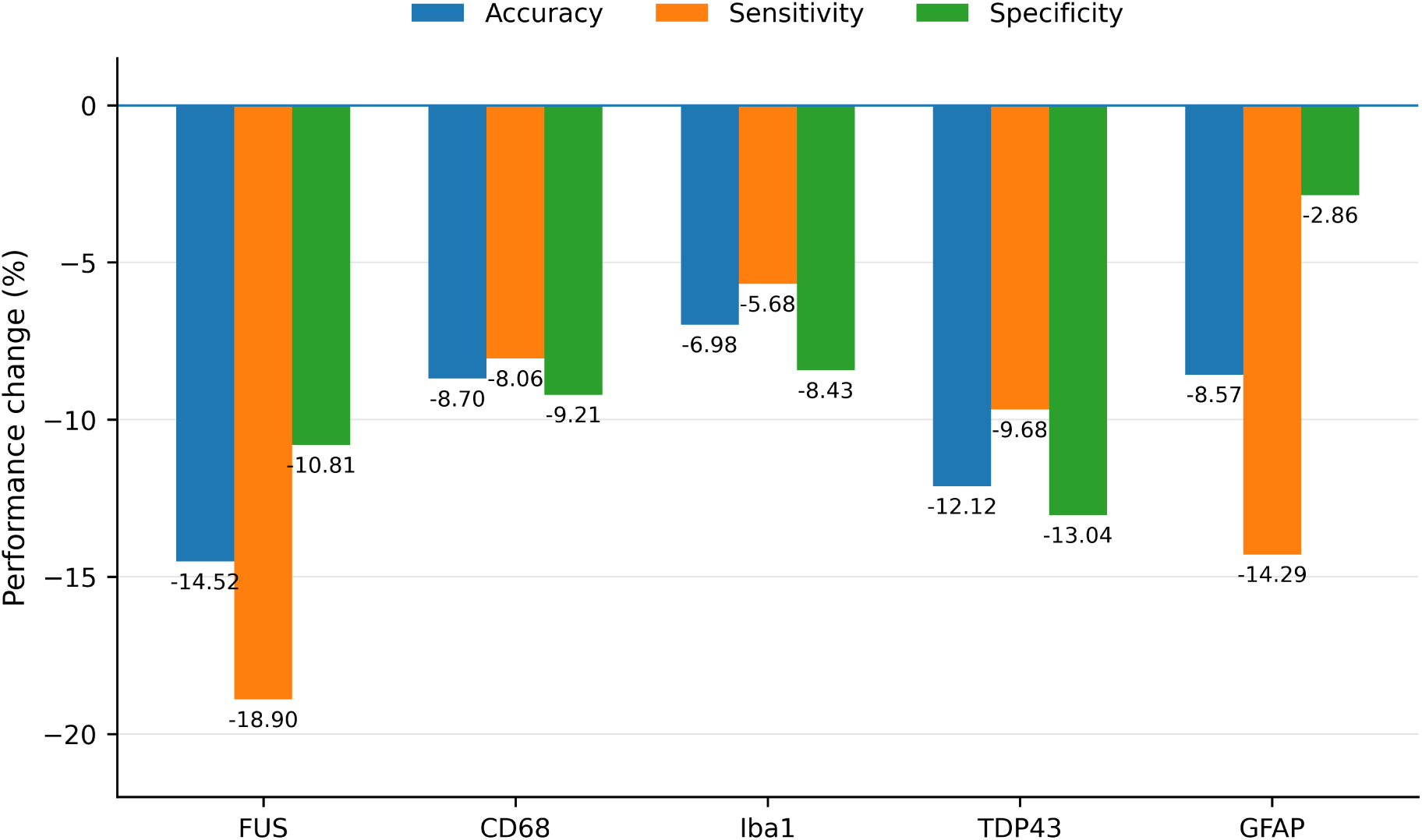
Effect of patient-isolated validation on random forest classification performance. Percentage change in accuracy, sensitivity and specificity of the 5-grouped cross-validation (5-g-CV) model relative to the conventional 3-fold cross-validation (3-CV) model for each biomarker. Negative values indicate lower performance under patient-isolated validation. Across all biomarkers, grouped cross-validation reduced overall classification performance, demonstrating that conventional cross-validation consistently overestimates model performance when patient-level data are not isolated during model evaluation.

**Figure 2.**
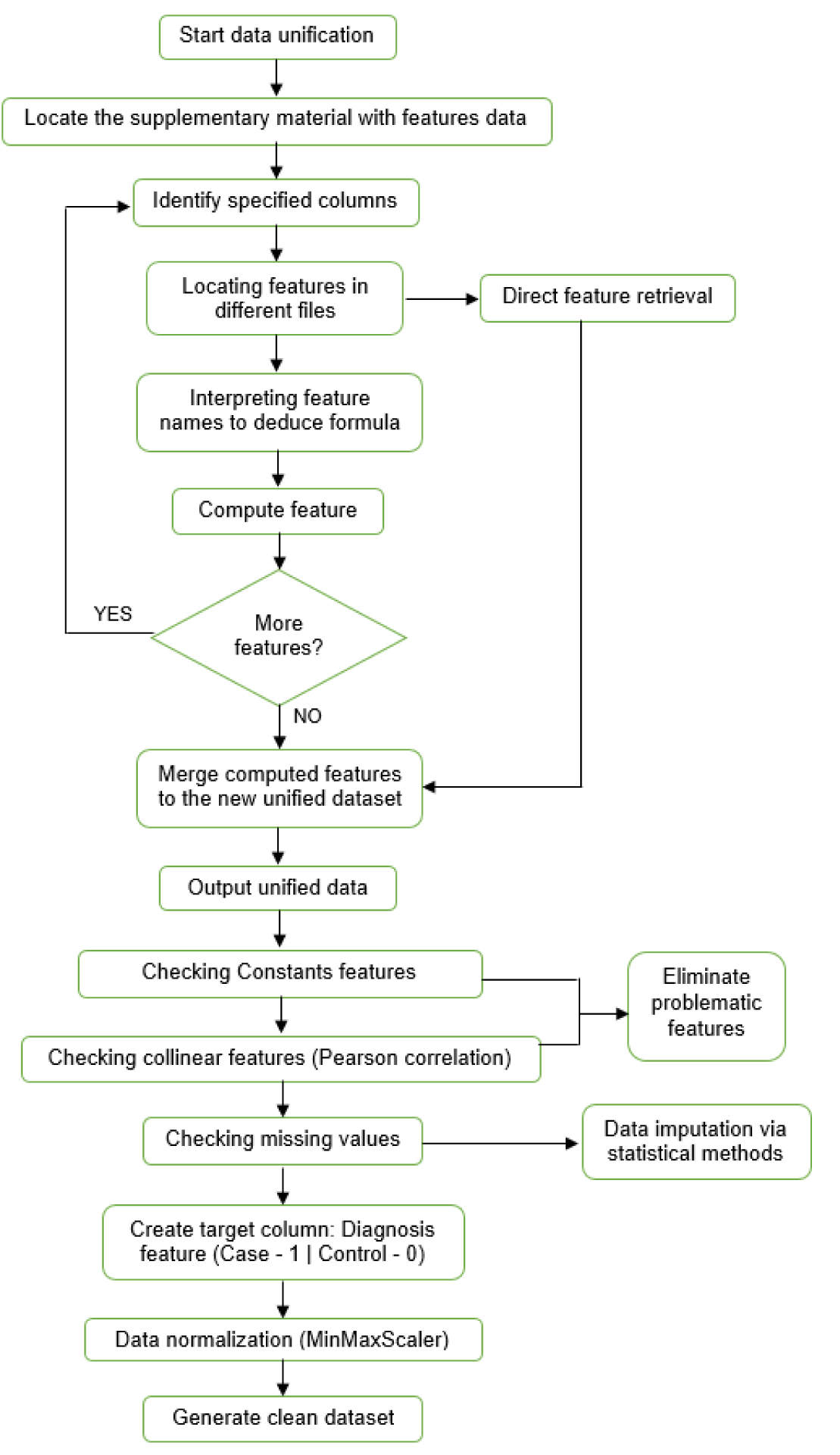
^1^Pre-processing pipeline used in this study.

**Figure 3.**
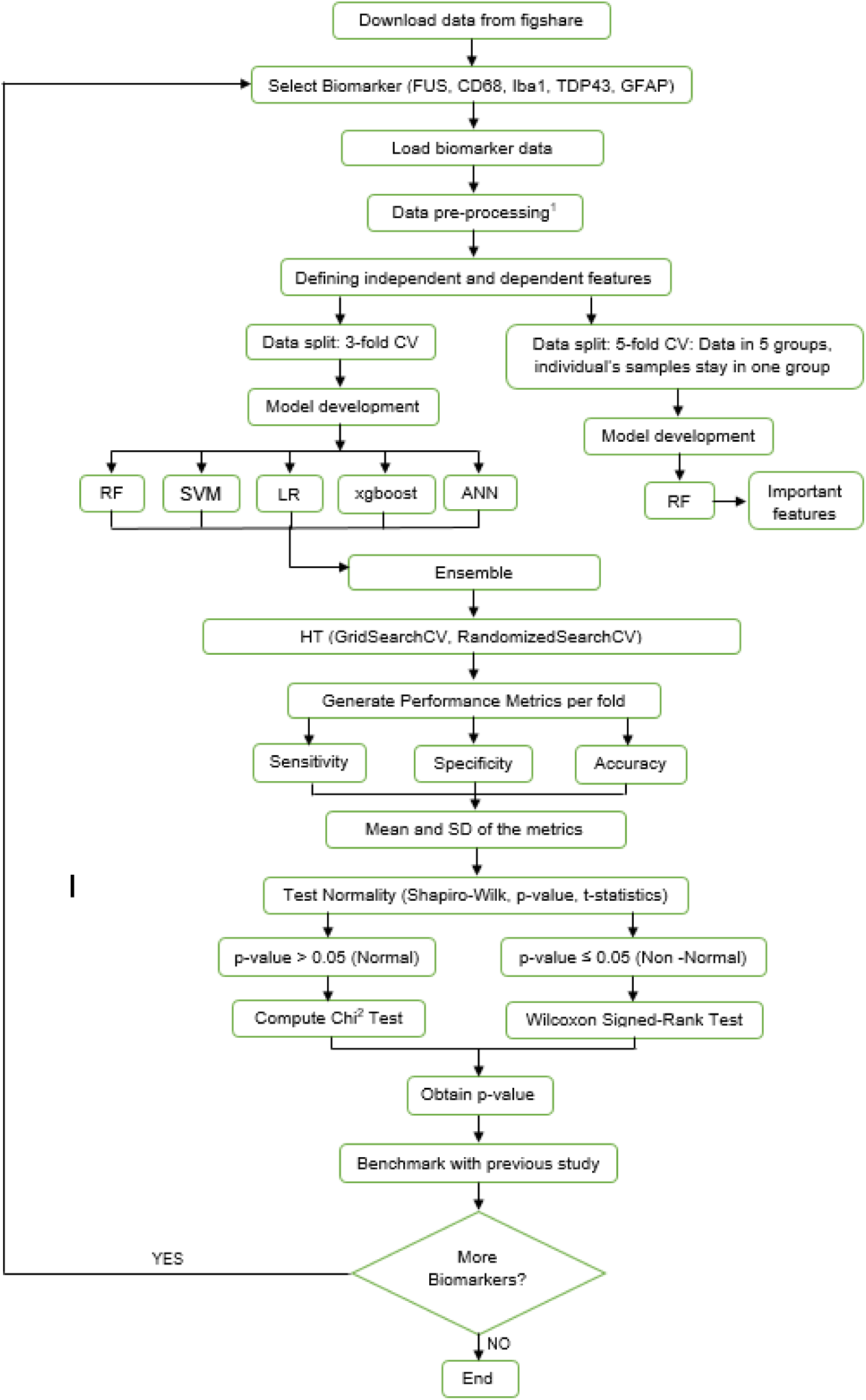
End-to-end machine learning workflow for building and evaluating models to identify C9-ALS. Abbreviations: Biomarkers used from the previous study: FUS - Fused in Sarcoma, CD68 - cluster of differentiation 68, Iba1 - ionised calcium-binding adapter molecule 1, TDP43 - TAR DNA-binding protein 43, and GFAP - glial fibrillary acidic protein. ML models implemented: RF – random forest, SVM – support vector machine, LR – logistic regression, XGBoost – extreme gradient boosting, ANN – artificial neural network. ML terminology: HT– hyperparameter tuning, CV – cross-validation, SD –standard deviation, stats – statistics

**Figure 4.**
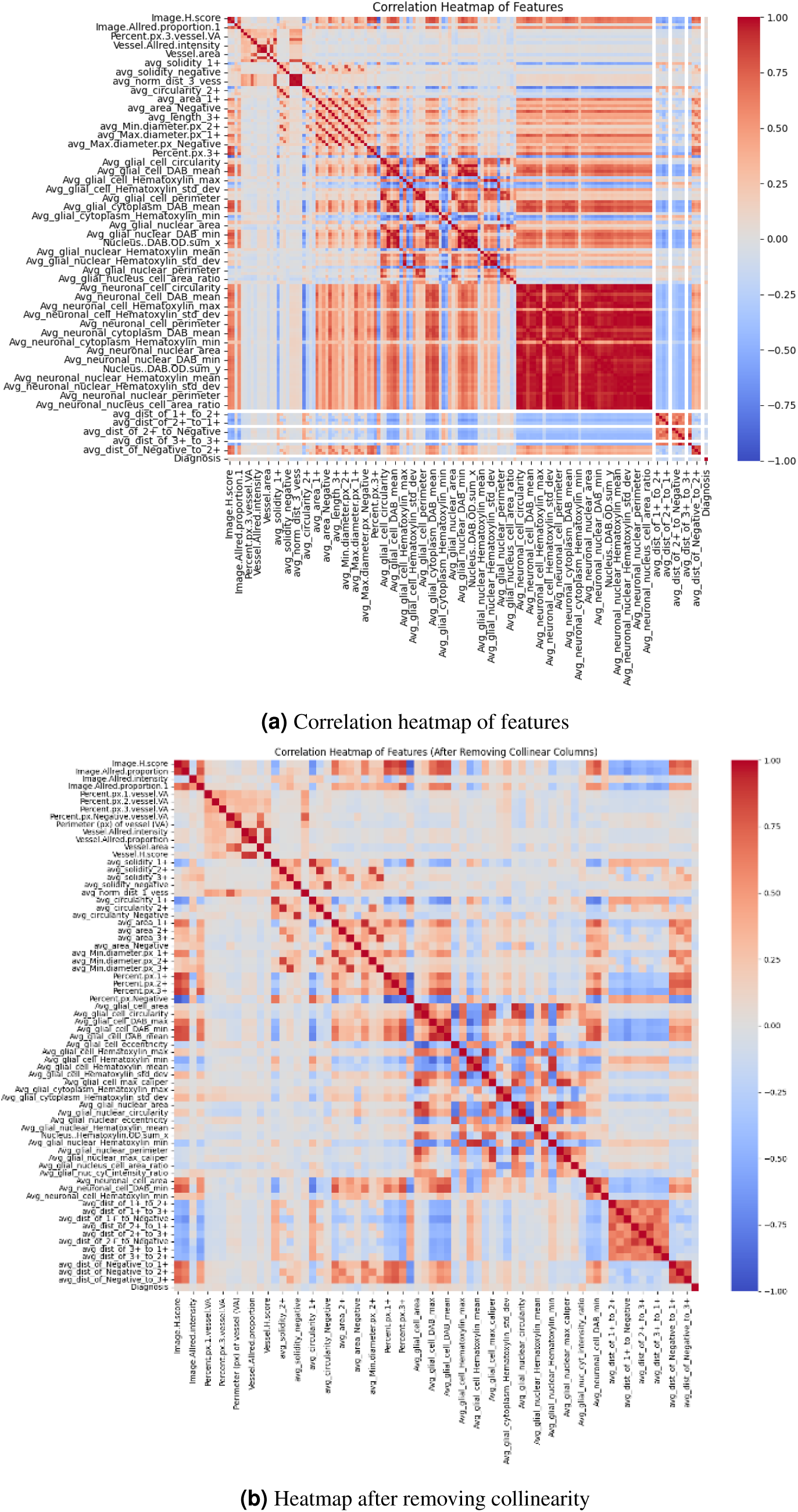
Heatmap of feature correlations of the FUS biomarker before and after removing collinearity.

**Figure 5.**
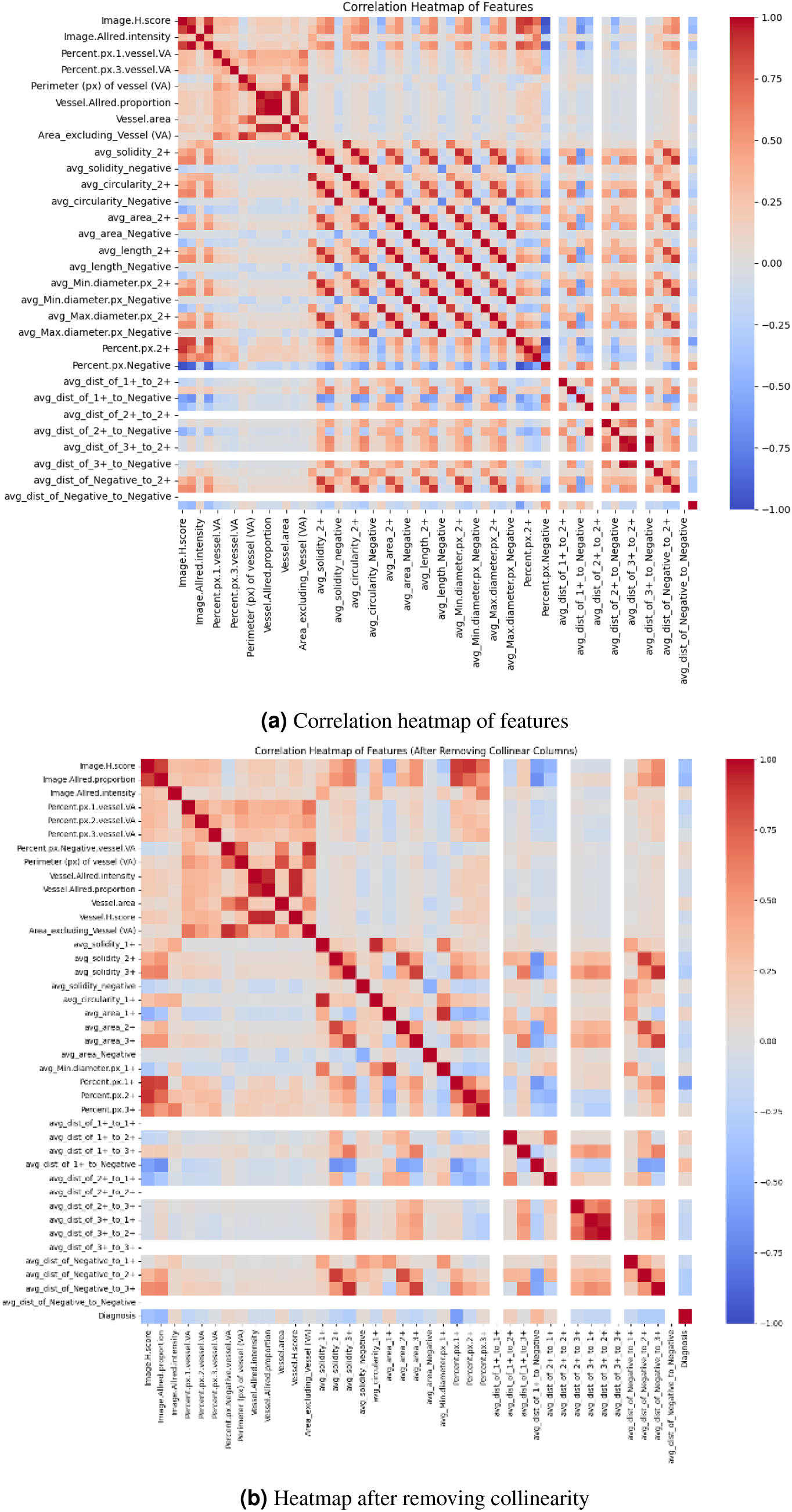
Heatmap of feature correlations of Iba1 biomarker before and after removing collinearity.

**Figure 6.**
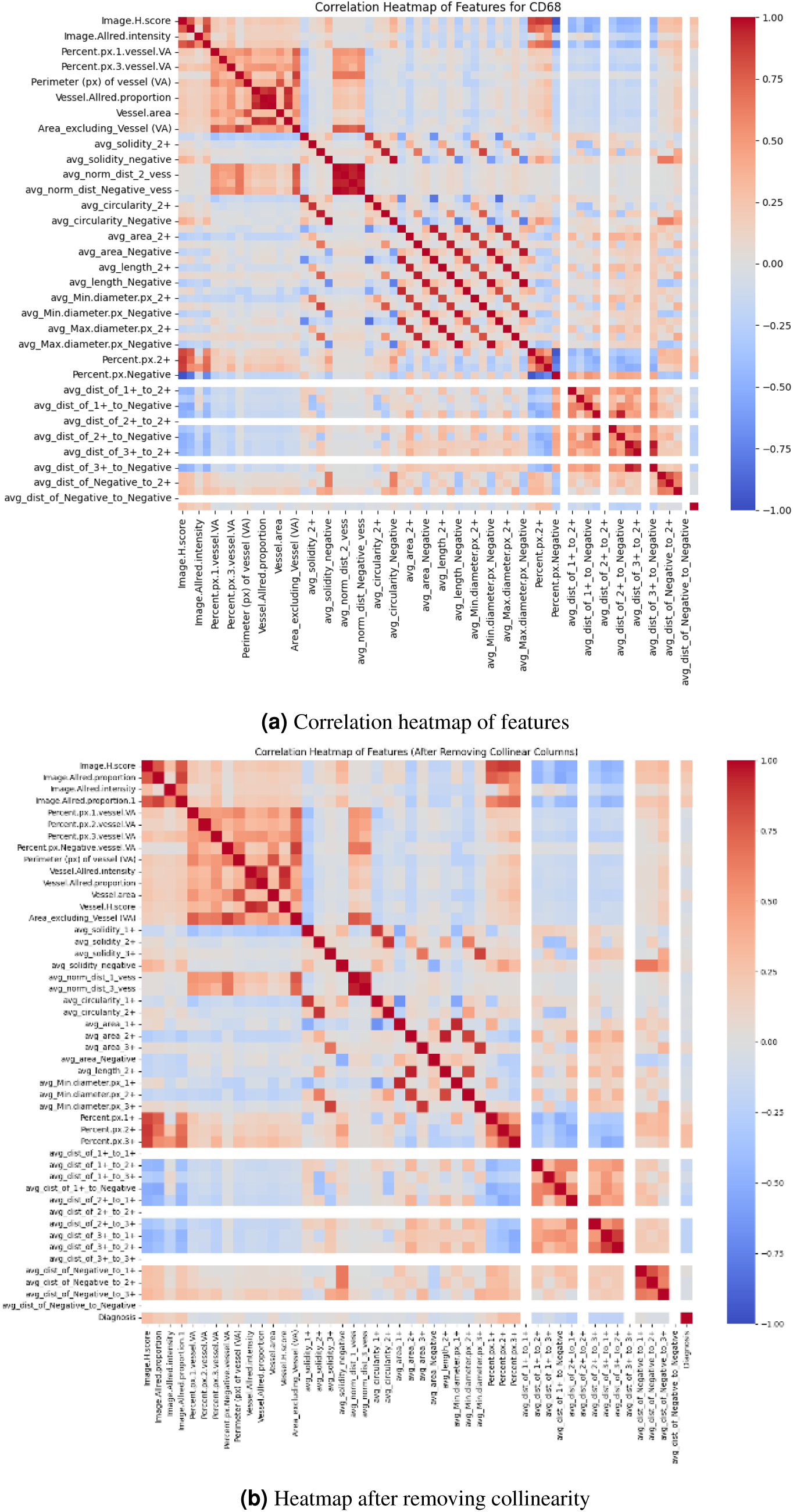
Heatmap of feature correlations of CD68 biomarker before and after removing collinearity.

**Figure 7.**
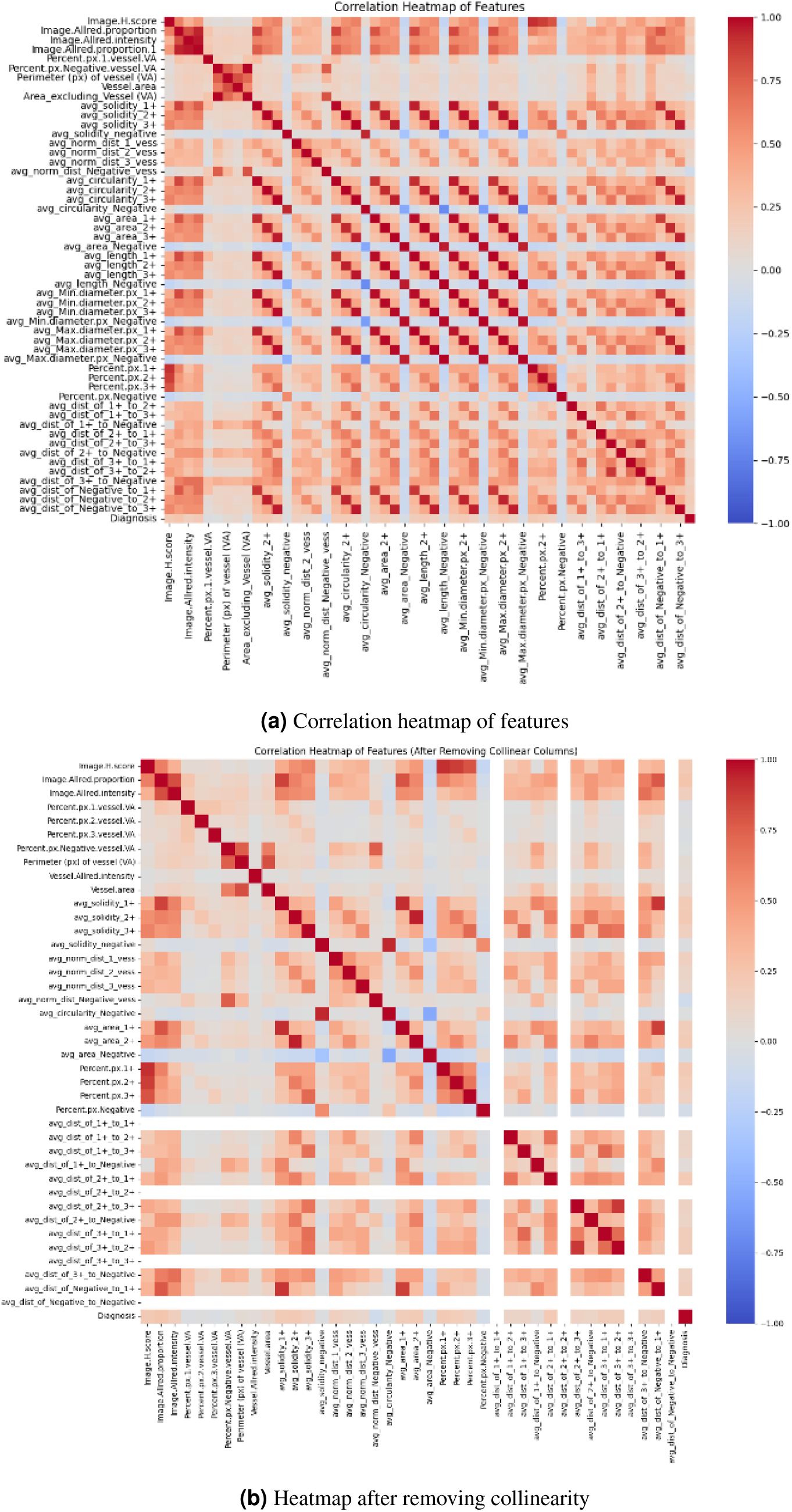
Heatmap of feature correlations of TDP43 biomarker before and after removing collinearity.

**Figure 8.**
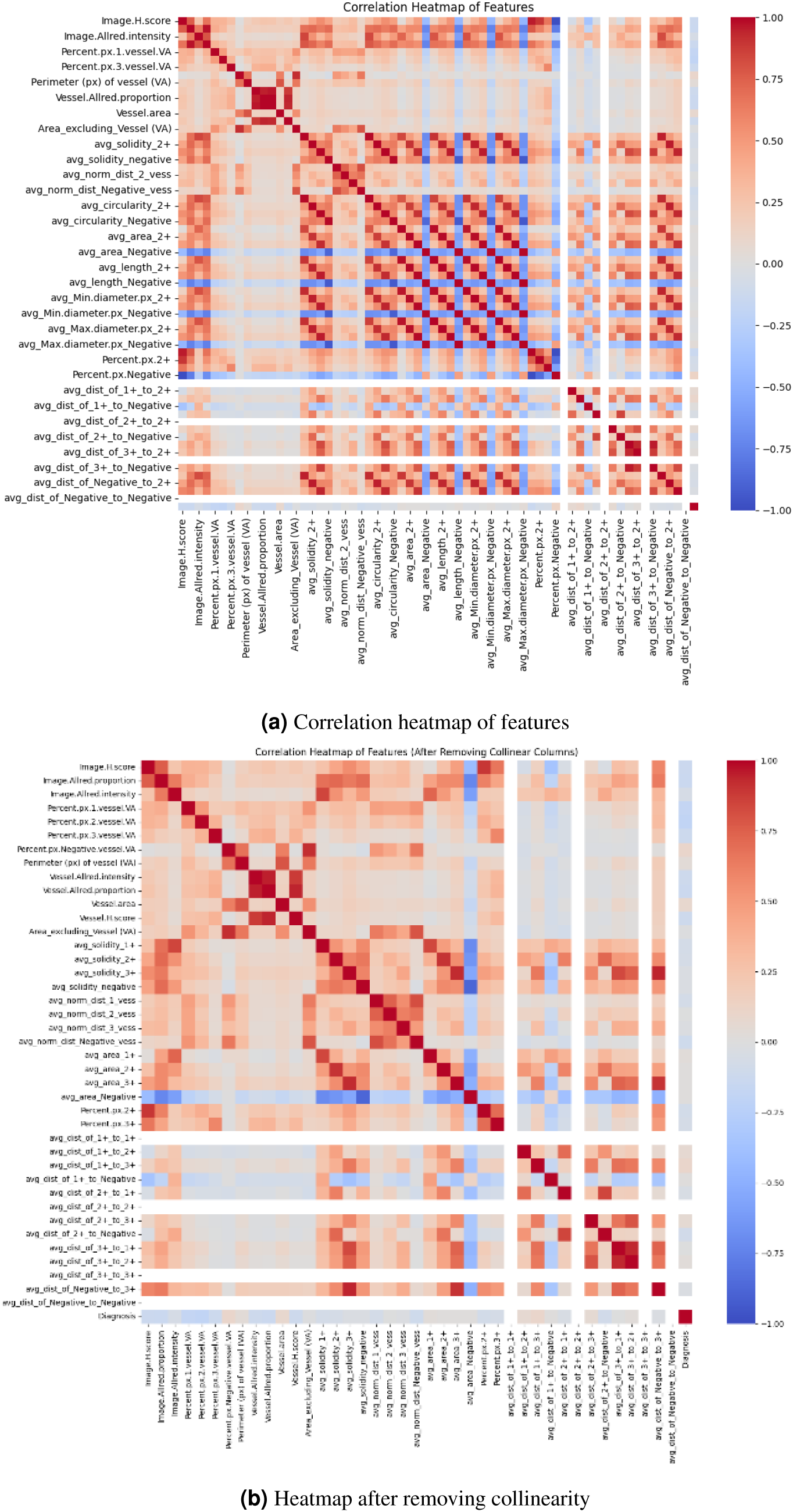
Heatmap of feature correlations of the GFAP biomarker before and after removing collinearity.

### Model training and validation results

Based on the main ML pipeline outlined in the Appendix (Figure 4), the performance of the complete set of models evaluated in the present study is reported separately for each biomarker in Tables 2–6. These tables provide a within-study comparison of random forest, SVM, logistic regression, XGBoost, ANN and the ensemble model. A separate comparison with the random forest results reported by Rifai *et al.*^11^ is presented subsequently, since the baseline study used a different experimental and validation framework.

**Table 2.** FUS classification results reported in the detailed model-performance table. Models evaluated using conventional 3-fold cross-validation (3-CV) are shown separately from patient-grouped 5-fold cross-validation (5-g-CV). Values are reported as mean *±* standard deviation as provided in the current manuscript.

| Model | Tuning | Validation | Leakage status | Sensitivity | Specificity | Accuracy |
| --- | --- | --- | --- | --- | --- | --- |
| Random Forest | No | 3-CV | Patient leakage | 0.69 ± 0.02 | 0.75 ± 0.04 | 0.72 ± 0.03 |
| Random Forest | Yes | 3-CV | Patient leakage | 0.73 ± 0.03 | 0.74 ± 0.03 | 0.73 ± 0.03 |
| SVM | No | 3-CV | Patient leakage | 0.71 ± 0.03 | 0.69 ± 0.04 | 0.70 ± 0.03 |
| SVM | Yes | 3-CV | Patient leakage | 0.71 ± 0.04 | 0.73 ± 0.04 | 0.72 ± 0.01 |
| Logistic Regression | No | 3-CV | Patient leakage | 0.69 ± 0.02 | 0.69 ± 0.04 | 0.69 ± 0.03 |
| Logistic Regression | Yes | 3-CV | Patient leakage | 0.73 ± 0.03 | 0.71 ± 0.02 | 0.72 ± 0.02 |
| XGBoost | No | 3-CV | Patient leakage | 0.68 ± 0.01 | 0.71 ± 0.04 | 0.70 ± 0.03 |
| XGBoost | Yes | 3-CV | Patient leakage | 0.708 ± 0.02 | 0.729 ± 0.02 | 0.72 ± 0.02 |
| ANN | No | 3-CV | Patient leakage | 0.519 ± 0.02 | 0.785 ± 0.02 | 0.652 ± 0.02 |
| ANN | Yes | 3-CV | Patient leakage | 0.73 ± 0.06 | 0.63 ± 0.06 | 0.68 ± 0.02 |
| Ensemble | No | 3-CV | Patient leakage | **0.76 ± 0.01** | 0.71 ± 0.04 | 0.73 ± 0.03 |
| Ensemble | Yes | 3-CV | Patient leakage | 0.75 ± 0.04 | 0.74 ± 0.04 | **0.74 ± 0.04** |
| Random Forest | Yes | 5-g-CV | Patient-isolated | 0.592 ± 0.10 | **0.66 ± 0.13** | 0.624 ± 0.04 |
| Random Forest | No | 5-g-CV | Patient-isolated | 0.64 ± 0.12 | 0.631 ± 0.13 | 0.633 ± 0.05 |
| Ensemble | Yes | 5-g-CV | Grouped outer split | **0.66 ± 0.08** | 0.65 ± 0.07 | **0.651 ± 0.04** |

**Table 3.**
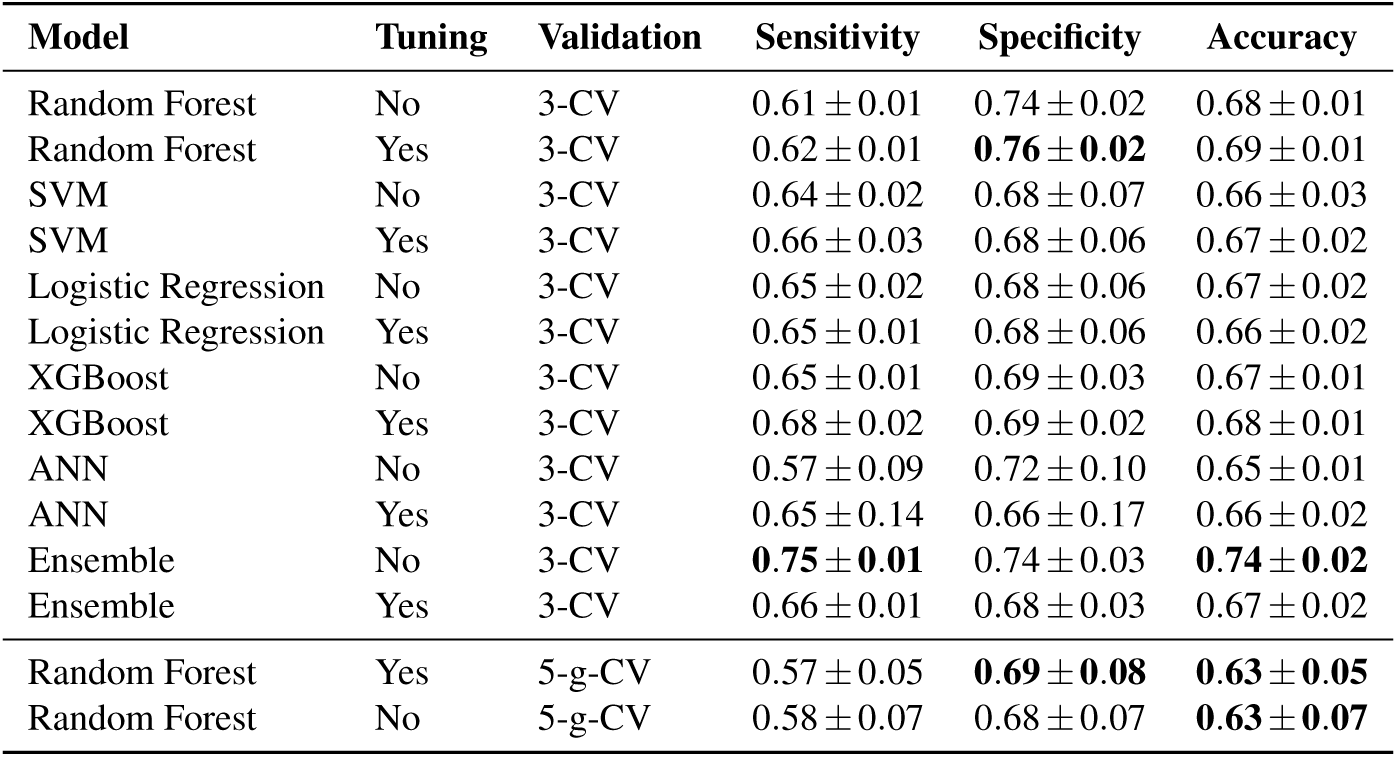
CD68 classification results reported in the detailed model-performance table. Models evaluated using conventional 3-fold cross-validation (3-CV) are shown separately from patient-grouped 5-fold cross-validation (5-g-CV). Values are reported as mean *±* standard deviation as provided in the current manuscript.

**Table 4.** Iba1 classification results reported in the detailed model-performance table. Models evaluated using conventional 3-fold cross-validation (3-CV) are shown separately from patient-grouped 5-fold cross-validation (5-g-CV). Values are reported as mean *±* standard deviation as provided in the current manuscript.

| Model | Tuning | Validation | Sensitivity | Specificity | Accuracy |
| --- | --- | --- | --- | --- | --- |
| Random Forest | No | 3-CV | $0.87 \pm 0.03$ | $0.82 \pm 0.02$ | $0.85 \pm 0.02$ |
| Random Forest | Yes | 3-CV | **$0.88 \pm 0.03$** | $0.83 \pm 0.01$ | **$0.86 \pm 0.01$** |
| SVM | No | 3-CV | $0.82 \pm 0.04$ | $0.82 \pm 0.04$ | $0.82 \pm 0.03$ |
| SVM | Yes | 3-CV | $0.83 \pm 0.04$ | $0.83 \pm 0.04$ | $0.83 \pm 0.03$ |
| Logistic Regression | No | 3-CV | $0.82 \pm 0.03$ | $0.82 \pm 0.04$ | $0.82 \pm 0.03$ |
| Logistic Regression | Yes | 3-CV | $0.83 \pm 0.04$ | $0.83 \pm 0.03$ | $0.83 \pm 0.03$ |
| XGBoost | No | 3-CV | $0.85 \pm 0.03$ | $0.83 \pm 0.02$ | $0.84 \pm 0.02$ |
| XGBoost | Yes | 3-CV | $0.86 \pm 0.03$ | $0.84 \pm 0.02$ | $0.85 \pm 0.02$ |
| ANN | No | 3-CV | $0.79 \pm 0.06$ | $0.82 \pm 0.05$ | $0.81 \pm 0.03$ |
| ANN | Yes | 3-CV | $0.82 \pm 0.05$ | $0.83 \pm 0.05$ | $0.83 \pm 0.03$ |
| Ensemble | No | 3-CV | $0.85 \pm 0.03$ | **$0.85 \pm 0.02$** | $0.85 \pm 0.02$ |
| Ensemble | Yes | 3-CV | $0.86 \pm 0.04$ | $0.84 \pm 0.02$ | $0.85 \pm 0.03$ |
| Random Forest | Yes | 5-g-CV | $0.83 \pm 0.06$ | $0.76 \pm 0.12$ | $0.80 \pm 0.06$ |
| Random Forest | No | 5-g-CV | $0.83 \pm 0.06$ | $0.76 \pm 0.11$ | $0.80 \pm 0.06$ |

**Table 5.** TDP43 classification results reported in the detailed model-performance table. Models evaluated using conventional 3-fold cross-validation (3-CV) are shown separately from patient-grouped 5-fold cross-validation (5-g-CV). Values are reported as mean *±* standard deviation as provided in the current manuscript.

| Model | Tuning | Validation | Sensitivity | Specificity | Accuracy |
| --- | --- | --- | --- | --- | --- |
| Random Forest | No | 3-CV | $0.60 \pm 0.01$ | $0.61 \pm 0.05$ | $0.60 \pm 0.30$ |
| Random Forest | Yes | 3-CV | **$0.62 \pm 0.03$** | $0.69 \pm 0.08$ | **$0.66 \pm 0.04$** |
| SVM | No | 3-CV | $0.47 \pm 0.05$ | $0.75 \pm 0.01$ | $0.61 \pm 0.03$ |
| SVM | Yes | 3-CV | $0.44 \pm 0.04$ | $0.81 \pm 0.04$ | $0.62 \pm 0.02$ |
| Logistic Regression | No | 3-CV | $0.47 \pm 0.06$ | $0.76 \pm 0.03$ | $0.61 \pm 0.03$ |
| Logistic Regression | Yes | 3-CV | $0.52 \pm 0.10$ | $0.71 \pm 0.06$ | $0.62 \pm 0.02$ |
| XGBoost | No | 3-CV | **$0.62 \pm 0.03$** | $0.57 \pm 0.07$ | $0.59 \pm 0.03$ |
| XGBoost | Yes | 3-CV | $0.48 \pm 0.07$ | $0.77 \pm 0.02$ | $0.62 \pm 0.04$ |
| ANN | No | 3-CV | $0.49 \pm 0.05$ | $0.71 \pm 0.06$ | $0.60 \pm 0.03$ |
| ANN | Yes | 3-CV | $0.36 \pm 0.11$ | **$0.85 \pm 0.09$** | $0.61 \pm 0.01$ |
| Ensemble | No | 3-CV | $0.56 \pm 0.07$ | $0.71 \pm 0.06$ | $0.63 \pm 0.04$ |
| Ensemble | Yes | 3-CV | $0.53 \pm 0.04$ | $0.73 \pm 0.02$ | $0.63 \pm 0.03$ |
| Random Forest | Yes | 5-g-CV | $0.56 \pm 0.09$ | $0.60 \pm 0.04$ | $0.58 \pm 0.03$ |
| Random Forest | No | 5-g-CV | $0.56 \pm 0.11$ | **$0.62 \pm 0.06$** | **$0.59 \pm 0.05$** |

**Table 6.** GFAP classification results reported in the detailed model-performance table. Models evaluated using conventional 3-fold cross-validation (3-CV) are shown separately from patient-grouped 5-fold cross-validation (5-g-CV). Values are reported as mean *±* standard deviation as provided in the current manuscript.

| Model | Tuning | Validation | Sensitivity | Specificity | Accuracy |
| --- | --- | --- | --- | --- | --- |
| Random Forest | No | 3-CV | $0.67 \pm 0.05$ | **$0.71 \pm 0.02$** | $0.69 \pm 0.03$ |
| Random Forest | Yes | 3-CV | $0.70 \pm 0.05$ | $0.70 \pm 0.03$ | **$0.70 \pm 0.02$** |
| SVM | No | 3-CV | $0.65 \pm 0.05$ | $0.63 \pm 0.01$ | $0.64 \pm 0.03$ |
| SVM | Yes | 3-CV | **$0.73 \pm 0.03$** | $0.62 \pm 0.02$ | $0.67 \pm 0.01$ |
| Logistic Regression | No | 3-CV | $0.68 \pm 0.04$ | $0.59 \pm 0.03$ | $0.64 \pm 0.03$ |
| Logistic Regression | Yes | 3-CV | $0.68 \pm 0.04$ | $0.65 \pm 0.01$ | $0.66 \pm 0.03$ |
| XGBoost | No | 3-CV | $0.67 \pm 0.04$ | $0.69 \pm 0.01$ | $0.68 \pm 0.02$ |
| XGBoost | Yes | 3-CV | $0.69 \pm 0.04$ | $0.69 \pm 0.02$ | $0.69 \pm 0.02$ |
| ANN | No | 3-CV | $0.62 \pm 0.05$ | $0.67 \pm 0.01$ | $0.65 \pm 0.03$ |
| ANN | Yes | 3-CV | $0.63 \pm 0.12$ | $0.66 \pm 0.07$ | $0.64 \pm 0.03$ |
| Ensemble | No | 3-CV | $0.70 \pm 0.06$ | $0.68 \pm 0.04$ | $0.69 \pm 0.02$ |
| Ensemble | Yes | 3-CV | **$0.73 \pm 0.05$** | $0.67 \pm 0.02$ | **$0.70 \pm 0.02$** |
| Random Forest | Yes | 5-g-CV | $0.60 \pm 0.10$ | $0.68 \pm 0.09$ | $0.64 \pm 0.08$ |
| Random Forest | No | 5-g-CV | $0.60 \pm 0.11$ | $0.68 \pm 0.12$ | $0.64 \pm 0.10$ |

The benchmark study by Rifai *et al.*^11^ reported a single performance estimate for each biomarker without measures of variability. In contrast, the present study reports the mean and standard deviation of performance metrics across the cross- validation folds, providing a measure of variability across folds. As accuracy was not reported by Rifai *et al.*, the benchmark accuracy values presented in this study were calculated from the performance results reported in the original publication to enable comparison across a consistent set of evaluation metrics.

Under conventional 3-CV, the tuned ensemble achieved the highest overall FUS accuracy (0.74 *±* 0.04), with sensitivity of 0.75 *±* 0.04 and specificity of 0.74 *±* 0.04. The untuned ensemble produced the highest sensitivity (0.76 *±* 0.01), whereas the highest specificity was observed for the untuned ANN (0.785 *±* 0.02). However, the latter showed markedly lower sensitivity (0.519 *±* 0.02), indicating an imbalanced classification profile. The tuned random forest remained competitive (0.73 *±* 0.03 accuracy; 0.73 *±* 0.03 sensitivity; 0.74 *±* 0.03 specificity) but was not the highest-performing 3-CV model. Because conventional 3-CV permits patient-level leakage, these results should be interpreted primarily as a benchmark against the original analysis rather than as estimates of performance on unseen patients.

Under 5-g-CV, random forest performance was lower than under conventional 3-CV. The untuned grouped random forest achieved an accuracy of 0.633 *±* 0.05, whereas the tuned grouped random forest achieved 0.624 *±* 0.04. These results suggest that hyperparameter tuning did not improve FUS performance under the reported grouped evaluation. However, the tuning procedure must first be confirmed to ensure that model selection was performed without using information from the held-out patient groups.

For CD68, performance differed considerably between models under conventional 3-CV. The untuned ensemble achieved the highest overall accuracy (0.74 *±* 0.02) and sensitivity (0.75 *±* 0.01), while maintaining a specificity of 0.74 *±* 0.03. The tuned random forest achieved the highest specificity (0.76 *±* 0.02), but with substantially lower sensitivity (0.62 *±* 0.01) and an accuracy of 0.69 *±* 0.01. The tuned XGBoost model produced a more balanced sensitivity and specificity of 0.68 *±* 0.02 and 0.69 *±* 0.02, respectively, but its overall accuracy (0.68 *±* 0.01) remained below that of the ensemble. Thus, among the reported conventional 3-CV models, the untuned ensemble provided the strongest overall CD68 classification performance.

Under patient-grouped 5-g-CV, random forest performance was lower than under conventional 3-CV. The tuned grouped random forest achieved a sensitivity of 0.57 *±* 0.05, specificity of 0.69 *±* 0.08, and accuracy of 0.63 *±* 0.05, while the untuned grouped model achieved 0.58 *±* 0.07, 0.68 *±* 0.07, and 0.63 *±* 0.07, respectively. The similar performance of the tuned and untuned grouped models suggests that hyperparameter optimisation did not materially improve CD68 classification under patient-isolated validation.

Under conventional 3-CV, the untuned ensemble—not the tuned ensemble, random forest or XGBoost—is the strongest CD68 model overall. Patient-isolated random forest performance is substantially lower, and hyperparameter tuning provides little or no benefit under grouped validation. However, the current manuscript contains a model-labelling inconsistency and appears to compare differently tuned random forest configurations when quantifying the effect of patient isolation.

Iba1 produced the strongest classification performance among the biomarkers examined under conventional 3-CV. The tuned random forest achieved the highest accuracy (0.86 *±* 0.01) and sensitivity (0.88 *±* 0.03), with a specificity of 0.83 *±* 0.01. The ensemble and XGBoost models produced closely comparable results. The untuned ensemble achieved the highest specificity (0.85 *±* 0.02), with sensitivity and accuracy of approximately 0.85, while the tuned XGBoost model achieved sensitivity of 0.86 *±* 0.03, specificity of 0.84 *±* 0.02, and accuracy of 0.85 *±* 0.02. Thus, random forest provided the highest overall 3-CV accuracy for Iba1, although several tree-based and ensemble approaches produced similar performance.

Compared with the random forest results reported by Rifai *et al.*, which achieved a sensitivity of 0.78 and a specificity of 0.84, the tuned 3-CV random forest increased sensitivity to 0.88 but produced a slightly lower specificity of 0.83. This corresponds to a 12.82% increase in sensitivity and a 1.19% decrease in specificity. However, this comparison should be interpreted cautiously because the conventional 3-CV evaluation permits observations from the same patient to occur in both training and validation folds.

Under patient-grouped 5-g-CV, random forest performance decreased. The reported grouped model achieved a sensitivity of approximately 0.83, a specificity of 0.76, and an accuracy of 0.80. Thus, although Iba1 remained the best-performing biomarker under the reported patient-isolated random forest analysis, its performance was lower than suggested by conventional 3-CV. The similar results obtained with tuned and untuned grouped random forest models also suggest that hyperparameter optimisation had little effect on Iba1 classification once patient-level isolation was imposed.

For TDP43, the tuned random forest provided the strongest overall performance among the models evaluated using conventional 3-CV, achieving a sensitivity of 0.62 *±* 0.03, specificity of 0.69 *±* 0.08, and accuracy of 0.66 *±* 0.04. Although the untuned XGBoost model achieved the same mean sensitivity (0.62 *±* 0.03), its lower specificity (0.57 *±* 0.07) resulted in an accuracy of only 0.59 *±* 0.03. Several other models achieved substantially higher specificity, including the tuned SVM (0.81 *±* 0.04) and tuned ANN (0.85 *±* 0.09), but these gains were accompanied by low sensitivities of 0.44 *±* 0.04 and 0.36 *±* 0.11, respectively. These models therefore exhibited pronounced class-specific imbalance rather than superior overall classification performance. Among the conventional 3-CV models, the tuned random forest provided the most favourable balance between sensitivity, specificity and accuracy.

Compared with the random forest results reported by Rifai *et al.*, which achieved a sensitivity of 0.51 and specificity of 0.64, the tuned 3-CV random forest increased sensitivity to 0.62 and specificity to 0.69. These changes correspond to increases of approximately 21.57% in sensitivity and 7.81% in specificity. Thus, the current statement that TDP43 performed more poorly than the baseline study is not supported by this direct random forest comparison. Nevertheless, the apparent improvement should be interpreted cautiously because the conventional 3-CV procedure does not isolate observations from the same patient between training and validation folds.

Under patient-grouped 5-g-CV, random forest performance decreased. The tuned grouped random forest achieved sensitivity of 0.56 *±* 0.09, specificity of 0.60 *±* 0.04, and accuracy of 0.58 *±* 0.03, whereas the untuned grouped model achieved sensitivity of 0.56 *±* 0.11, specificity of 0.62 *±* 0.06, and accuracy of 0.59 *±* 0.05. The grouped results therefore provide substantially weaker evidence of TDP43-based discrimination than the conventional 3-CV results and indicate that hyperparameter optimisation did not improve performance under the reported patient-isolated evaluation.

For GFAP, no single model achieved the highest value across all conventional 3-CV performance metrics. The tuned random forest and tuned ensemble shared the highest mean accuracy (0.70 *±* 0.02). The tuned random forest produced balanced sensitivity and specificity of 0.70 *±* 0.05 and 0.70 *±* 0.03, respectively. In comparison, the tuned ensemble achieved higher sensitivity (0.73 *±* 0.05) but lower specificity (0.67 *±* 0.02). The tuned SVM also achieved the highest mean sensitivity (0.73 *±* 0.03), but this was accompanied by substantially lower specificity (0.62 *±* 0.02) and an accuracy of 0.67 *±* 0.01. The highest specificity was obtained with the untuned random forest (0.71 *±* 0.02), which achieved a sensitivity of 0.67 *±* 0.05 and accuracy of 0.69 *±* 0.03. Consequently, the tuned random forest provided the most balanced conventional 3-CV classification profile, although the tuned ensemble achieved equivalent overall accuracy.

The random forest results also differed from those reported by Rifai *et al.*, whose GFAP model achieved a sensitivity of 0.79 and specificity of 0.53. The tuned 3-CV random forest in the present study achieved a lower sensitivity of 0.7 but a substantially higher specificity of 0.70, corresponding to an 11.39% reduction in sensitivity and a 32.08% increase in specificity. Thus, the present model did not uniformly improve GFAP classification relative to the baseline; rather, it produced a more balanced sensitivity–specificity profile. This comparison should nevertheless be interpreted cautiously because the conventional 3-CV analysis does not isolate observations belonging to the same patient between training and validation folds.

Under patient-grouped 5-g-CV, random forest performance decreased further. Both the tuned and untuned grouped models achieved mean sensitivity of 0.6, specificity of 0.68, and accuracy of 0.64, although the reported variability differed between configurations. These results indicate that the apparent balance achieved under conventional 3-CV was reduced when patient-level isolation was introduced, particularly through a decrease in sensitivity. Hyperparameter optimisation did not improve the mean GFAP classification performance under the reported grouped evaluation.

Across all five biomarkers, random forest performance decreased when conventional 3-CV was replaced by patient-grouped 5-g-CV (Figure 1 and Table 14 in the Appendix). Accuracy decreased by 6.98–14.52%, sensitivity by 5.68–18.9%, and specificity by 2.86–13.04%, depending on the biomarker. The largest reductions in accuracy and sensitivity were observed for FUS, whereas the largest reduction in specificity occurred for TDP43. Iba1 showed the smallest reductions in accuracy and sensitivity and remained the strongest-performing biomarker under patient-grouped evaluation. These results indicate that estimated classification performance is sensitive to the validation strategy and that conventional observation-level CV provides more optimistic estimates than evaluation in which patients are isolated between training and test folds. However, because hyperparameter optimisation was not nested within the patient-grouped evaluation, the 5-g-CV results should not be interpreted as estimates from a fully leakage-free modelling pipeline. A direct comparison between the random forest performance reported by Rifai *et al.*^11^ and the random forest results obtained in the present study under conventional 3-CV and patient-grouped 5-g-CV is provided in Appendix Table 11.

### Feature Importance

Feature importance differed across the five biomarker-specific random forest models (Table 7). For FUS, the average glial nuclear-to-cytoplasmic intensity ratio had the highest mean Gini importance (0.023). Importance was distributed relatively evenly across the remaining top-ranked features, with no individual feature accounting for a large proportion of the model’s total feature importance.

**Table 7.** Feature importance for each biomarker computed using a random forest model, based on the mean decrease in the Gini impurity.

| Rank | Features | Importance | Biomarker |
| --- | --- | --- | --- |
| 1 | Average glial nuclear cytoplasmic intensity ratio | 0.023 | FUS |
| 2 | Average area for the class 3+ | 0.018 |  |
| 3 | Average minimum distance (px) for class 3 | 0.017 |  |
| 4 | Percent of super pixels that are class–Negative | 0.016 |  |
| 5 | Average solidity for class 3+ | 0.016 |  |
| 6 | Average glial cell Hematoxylin std dev | 0.016 |  |
| 7 | Average glial nuclear DAB max | 0.014 |  |
| 8 | Average glial nuclear DAB std dev | 0.014 |  |
| 9 | Average length for class 3+ | 0.014 |  |
| 10 | Average distance from 2+ to Negative | 0.014 |  |
| 1 | Percentage of super pixels that are class 1+ | 0.126 | Iba1 |
| 2 | Average distance of class 1+ to Negative | 0.068 |  |
| 3 | Average distance of 1+ to 2+ | 0.060 |  |
| 4 | Percentage of superpixels that are class Negative | 0.050 |  |
| 5 | Image H-score | 0.041 |  |
| 6 | Average distance from 1+ to 3+ | 0.028 |  |
| 7 | Average distance from Negative to 2+ | 0.027 |  |
| 8 | Percentage of super pixels that are class 3+ | 0.026 |  |
| 9 | Average distance from 3+ to 2+ | 0.025 |  |
| 10 | Average distance from 3+ to 1+ | 0.024 |  |
| 1 | Percentage of super pixels that are class 3+ | 0.042 | CD68 |
| 2 | Average distance of class 2+ to Negative | 0.040 |  |
| 3 | Percentage of super pixels that are class 2+ | 0.038 |  |
| 4 | Average distance of class 2+ to 1+ | 0.033 |  |
| 5 | Average distance of class 3+ to 1+ | 0.032 |  |
| 6 | Image H-score | 0.029 |  |
| 7 | Average distance of class 3+ to Negative | 0.027 |  |
| 8 | Average maximum diameter (px) of class 1+ | 0.025 |  |
| 9 | Average length of class 3+ | 0.025 |  |
| 10 | Average distance of class 3+ to 2+ | 0.025 |  |
| 1 | Average minimum diameter (px) of class Negative | 0.060 | TDP43 |
| 2 | Average maximum diameter (px) of class Negative | 0.056 |  |
| 3 | Average solidity of class Negative | 0.055 |  |
| 4 | Average area of class negative | 0.055 |  |
| 5 | Average length of class negative | 0.054 |  |
| 6 | Average circularity of class negative | 0.053 |  |
| 7 | Area excluding vessel (VA) | 0.049 |  |
| 8 | Average normalised distance of class negative in vessel | 0.047 |  |
| 9 | Vessel area | 0.042 |  |
| 10 | Average distance of class 1+ to Negative | 0.036 |  |
| 1 | Average minimum diameter (px) of class Negative | 0.044 | GFAP |
| 2 | Average maximum diameter (px) of class Negative | 0.044 |  |
| 3 | Average area of class negative | 0.043 |  |
| 4 | Average length of class negative | 0.042 |  |
| 5 | Average solidity of class negative | 0.041 |  |
| 6 | Average circularity of class negative | 0.038 |  |
| 7 | Average maximum diameter (px) of class 1+ | 0.032 |  |
| 8 | Average area of class 1+ | 0.031 |  |
| 9 | Average circularity of class 1+ | 0.028 |  |
| 10 | Image H-Score | 0.027 |  |

For Iba1, the percentage of class 1+ superpixels was the highest-ranked feature, with a mean importance of 0.126, whereas each of the remaining nine highest-ranked features had importance values below 0.068. For CD68, the percentage of class 3+ superpixels had the highest importance (0.042). TDP43 and GFAP shared the same highest-ranked feature, the average minimum diameter of negative-class superpixels, with mean importance values of 0.060 and 0.044, respectively.

Overall, the biomarker-specific models exhibited different feature-importance profiles, with the strongest individual feature contribution observed for Iba1. Nevertheless, importance was distributed across multiple features for all five biomarkers, indicating that classification was not determined by a single dominant feature.

## 3 Discussion

This study builds on the digital pathology framework established by Rifai *et al.*^11^, while addressing a different computational objective. Whereas the baseline study primarily investigated whether quantitative pathological features could characterise C9orf72-associated disease status and phenotype, the present study examined the robustness of the machine learning framework used to evaluate these features. The analysis reconstructed the biomarker-specific feature datasets, extended the classification framework beyond random forest to SVM, logistic regression, XGBoost, ANN and ensemble approaches, investigated preprocessing and hyperparameter optimisation, introduced patient-grouped evaluation, and examined biomarker-specific feature importance.

Exploratory analysis identified constant features and substantial multicollinearity within the reconstructed datasets. Removing constant or highly correlated features did not consistently improve classification performance and, in several cases, resulted in modest reductions in sensitivity, specificity or accuracy. The features specified in the baseline study were therefore retained for the subsequent analyses. However, the absence of a performance benefit from feature removal should not be interpreted as evidence that all variables contribute independently to classification. In particular, correlated predictors may contain overlapping information, an issue that also affects interpretation of the subsequent feature-importance analysis.

Hyperparameter optimisation produced relatively modest and model-dependent changes in performance. Improvements were not uniform across biomarkers or evaluation metrics; increases in one metric were sometimes accompanied by reductions in another. For example, optimisation of the random forest model for Iba1 increased sensitivity while slightly reducing specificity. These findings indicate that hyperparameter optimisation can alter the balance between class-specific performance measures rather than uniformly improving classification. The effect of optimisation should therefore be assessed across sensitivity, specificity and accuracy rather than using a single performance metric.

Extending the analysis beyond random forest further showed that no single algorithm consistently achieved the highest performance across all biomarkers and evaluation metrics. For several biomarkers, models other than random forest achieved higher sensitivity, specificity or accuracy under conventional 3-CV. The relative performance of the classifiers was therefore both biomarker- and metric-dependent, and the results do not support the selection of random forest or the ensemble as a universally superior classifier. However, these comparisons require caution because the non-random forest models were evaluated using conventional 3-CV and have not been assessed using the same patient-grouped validation framework. Their higher observed performance therefore cannot currently be interpreted as evidence of superior generalisation to unseen patients.

The comparison between conventional 3-CV and patient-grouped 5-g-CV provides further evidence of the importance of validation design. Random forest performance decreased under 5-g-CV for every biomarker and across sensitivity, specificity and accuracy (Figure 1; Table 14). The magnitude of the reduction varied across biomarkers and metrics, indicating that the effect of patient isolation was not uniform. Accuracy decreased by 6.98–14.52%, sensitivity by 5.68–18.90%, and specificity by 2.86–13.04%. FUS showed the largest reductions in accuracy and sensitivity, whereas TDP43 showed the largest reduction in specificity. In contrast, Iba1 showed the smallest reductions in accuracy and sensitivity and remained the strongest-performing random forest biomarker under patient-grouped evaluation. These findings demonstrate that allowing observations from the same individual to contribute to different cross-validation folds can produce more optimistic estimates of predictive performance.

Comparison with the baseline study further illustrates the influence of validation design (Appendix Table 11). Under conventional 3-CV, the random forest models in the present study generally achieved performance comparable to or higher than that reported by Rifai *et al.*^11^, although the magnitude and direction of the differences varied across biomarkers and metrics. However, these apparent improvements were reduced or reversed for several biomarkers when random forest was evaluated using patient-grouped 5-g-CV. This reinforces the observation that performance estimates obtained when observations from the same individual can contribute to different folds may be optimistic.

Importantly, the relative predictive value of the biomarkers showed greater consistency than the absolute performance estimates. Iba1 remained the strongest random forest biomarker under patient-grouped evaluation, consistent with the strong predictive performance reported for Iba1 in the baseline study. This suggests that some of the pathological signal identified by Rifai *et al.*^11^ is retained when patient-level separation is introduced, despite the reduction in absolute classification performance. Nevertheless, differences between the experimental and validation procedures mean that these results should not be interpreted as a strict like-for-like benchmark.

Feature-importance analysis indicated that disease-status classification was supported by multiple pathological features rather than being dominated by a single measurement. This was particularly evident for FUS, for which the highest-ranked feature had a Gini importance of only 0.023. Although importance was distributed across multiple features for all biomarkers, the profiles were not identical. Iba1 showed the strongest individual contribution, with the percentage of class 1+ superpixels reaching an importance of 0.126, suggesting a greater concentration of predictive information in this feature than was observed for the other biomarkers.

These findings are broadly consistent with Rifai *et al.*^11^, whose leave-one-feature-out analysis of a random forest incorporating features across the biomarker panel found that no individual feature significantly affected predictive performance when removed. However, the two analyses address different questions. Rifai *et al.* assessed feature contribution across the combined pathological feature space before grouping the identified features into pathological categories, whereas the present study evaluates feature importance independently within each biomarker-specific model. The present analysis therefore provides complementary information on how predictive information is distributed within individual pathological markers rather than reproducing the global feature-ranking analysis of the baseline study.

The feature-importance results should nevertheless be interpreted cautiously. Gini-based importance is model-dependent and can be affected by correlation between predictors. As highly correlated features were identified in the present datasets and retained because their removal did not improve model performance, some predictive information is likely to be shared among related variables. This can distribute importance across correlated features, making the ranking of individual variables less stable and potentially less representative of their underlying biological relevance. Furthermore, the current analysis reports mean importance values without quantifying the variability or stability of feature rankings across different patient partitions. Accordingly, the reported importance scores are best interpreted as exploratory patterns within each biomarker rather than as definitive evidence that individual pathological measurements are dominant disease drivers.

The statistical analysis based on fold-level performance estimates should also be interpreted cautiously. With only three observations under 3-CV and five under 5-g-CV, normality testing and subsequent inferential tests have very limited statistical power. The repeated or non-significant *p*-values obtained with the Wilcoxon signed-rank test therefore provide limited information about differences in model performance. More robust statistical assessment would require repeated patient-level cross-validation, providing a larger distribution of performance estimates from which uncertainty and model differences could be assessed more reliably.

A substantial component of this study involved reconstructing the feature sets required to reproduce and extend the baseline analysis. The publicly available data consisted of QuPath-derived pathological measurements rather than the final engineered features used for model training, while the supplementary material of Rifai *et al.*^11^ listed the derived features without fully specifying their computational reconstruction. Consequently, the required attributes had to be identified, interpreted and reconstructed from the available outputs for each biomarker. The resulting five structured, ML-ready biomarker datasets represent an additional output of the present work and provide a reproducible basis for subsequent computational investigation of this cohort.

### Limitations

Several limitations should be considered when interpreting these findings. First, the study was based on a relatively small cohort of 10 C9orf72-ALS cases and 10 controls. Although multiple tissue regions and observations were available per individual, these do not represent independent subjects. The use of grouped cross-validation prevented observations from the same individual from being shared between training and validation sets, but cannot compensate for the limited number of independent cases. The small cohort therefore restricts statistical power, increases uncertainty in model and hyperparameter selection, and limits the generalisability of the reported performance estimates.

Second, no independent external cohort was available for validation. All models were developed and evaluated using data derived from the same cohort, and their performance therefore requires confirmation in independent tissue collections. Furthermore, the disease cohort was restricted to C9orf72-associated ALS. Given the substantial genetic, pathological and clinical heterogeneity of ALS, the findings cannot be assumed to generalise to sporadic ALS, other genetic forms of ALS, or the wider ALS-frontotemporal spectrum.

The analysis also relies on quantitative pathological features previously extracted from post-mortem tissue using the digital pathology pipeline described by Rifai *et al.*^11^. Limitations arising from tissue sampling, ROI selection, staining, image segmentation and feature extraction may therefore propagate into the machine learning analysis. Although the underlying data and analysis resources were made publicly available, reconstruction of an ML-ready tabular dataset and interpretation of some derived variables required additional processing. This highlights the importance of providing clearly documented and directly reusable datasets alongside computational pathology studies to support reproducibility and independent methodological evaluation.

The collinearity-reduction analysis used a simple pair-order rule to remove one feature from each highly correlated pair rather than selecting variables based on biological relevance or predictive contribution. The resulting comparison therefore evaluates the effect of reducing redundancy rather than identifying an optimal non-collinear feature subset.

Feature-importance results should also be interpreted cautiously. Highly correlated features were identified in the dataset and were retained because their removal did not improve classification performance. In the presence of correlated predictors, random forest models may distribute importance across variables containing overlapping information, potentially reducing or destabilising the apparent importance of individual features. In addition, Gini-based feature importance is model-dependent and may be influenced by properties of the predictor variables. The resulting rankings should therefore be interpreted as relative contributions to classification within the fitted models rather than as direct evidence of the biological importance of individual pathological features. These considerations also support interpretation at the level of broader feature families rather than relying solely on individual feature rankings.

Hyperparameter optimisation was performed using accuracy as the optimisation criterion. Although accuracy provides an intuitive measure of overall classification performance and the dataset was balanced at the subject level, it does not capture class-specific performance or the trade-off between sensitivity and specificity. Model selection based on alternative or complementary metrics, such as balanced accuracy or the area under the receiver operating characteristic curve (AUC), may therefore result in different hyperparameter configurations. This consideration is particularly relevant given the small number of independent subjects, which may increase variability in hyperparameter selection. Although patient-grouped cross-validation prevented samples from the same individual from appearing simultaneously in the outer training and validation sets, hyperparameter optimisation was not implemented using a fully nested patient-grouped cross-validation framework. For the individual models, hyperparameters were selected using cross-validation on the dataset before subsequent performance evaluation, meaning that model selection and performance estimation were not fully independent. In addition, some data-dependent preprocessing operations were performed before the hyperparameter-search cross-validation rather than within the corresponding training folds. These factors may introduce optimistic bias into the reported tuned-model performance and should therefore be considered when interpreting the results.

Finally, the analysis is based exclusively on post-mortem tissue and should not be interpreted as demonstrating diagnostic or prognostic performance in living patients. Rather, the models provide a computational framework for investigating disease-associated pathological patterns in C9orf72-ALS. Validation in larger, independent and more diverse cohorts will be required before broader biological or translational conclusions can be established.

## 4 Methods

### 4.1 Original Dataset

The dataset utilised in the present study was taken from the research presented in^11^, which extensively analysed thousands of postmortem brain tissue images, generated through detailed imaging and digital processing of distinct tissue samples to identify features linked to glial activation, such as Iba1 and CD68, and protein misfolding, like FUS and TDP-43. Rifai *et al.* provided a systematic analysis of IHC markers related to intensity, morphology, and spatial patterns, as well as clinical and genetic information from 10 C9-ALS and 10 C9-ALS-FTSD patients and age- and sex-matched controls, with 12 samples from each of the four brain regions, Brodmann areas (BAs) BA4, BA39, BA44, and BA46, with a total of 48 images per individual. Each BA provided six samples from both grey and white matter, with three images obtained from non-vascular-adjacent (NVA) regions and three from vascular-adjacent (VA) regions. The dataset was balanced by assigning equal numbers of samples to C9-ALS cases and control groups across brain regions and tissue types, providing a well-matched comparison for analysis. Clinical and neuropathological characteristics of the C9-ALS and control cohorts, including disease duration, region of onset, cognitive assessment (ECAS), and availability of ALSFRS data, are provided in Appendix A, Table 17.

### 4.2 Data Integration and Curation

The dataset obtained from the online repository Figshare^11^ included data on five biomarkers: FUS, Iba1, CD68, GFAP, and TDP43. Each biomarker dataset was represented by a separate CSV file containing features extracted from post-mortem tissues^11^. For this study, the entire tabular dataset per biomarker was utilised, without directly analysing the image data, mirroring the work of Rifai *et al.*.

A total of 30 files were downloaded, six for each biomarker: FUS, CD68, Iba1, TDP43 and GFAP. The files contained quantitative digital pathology measurements of intensity, morphology and spatial patterns exported from QuPath, together with common identifiers including image name, class and ROI. However, the engineered features used to train the models reported by Rifai *et al.*^11^ were not provided directly in an ML-ready format.

To reconstruct the feature sets used in the original study, the available measurements were matched to the attributes reported in the supplementary material of Rifai *et al.*^11^ Table 10). Where required, features were calculated as means, percentages or other statistical summaries of the underlying measurements. The reconstructed features were organised into five biomarker-specific tabular datasets corresponding to FUS, CD68, Iba1, TDP43 and GFAP. The generation of these structured, ML-ready datasets represents an output of the present study and provides a reproducible basis for subsequent computational analyses. These datasets were then subjected to the pre-processing, model training and evaluation procedures described below.

For FUS, the cleaned dataset includes 960 rows and 149 features, while each of the other four biomarkers has 960 rows and 65 features. The target column, diagnosis, was assigned binary values: 0 for controls and 1 for cases. The final dataset still maintained a perfectly balanced class distribution. Detailed information on the dataset files for each biomarker and their features is provided in Table 10.

#### 4.2.1 Pre-processing of the generated datasets

The next stage focused on preparing each of the five biomarker-specific datasets for model training. A detailed exploratory data analysis (EDA) was conducted independently for the FUS, CD68, Iba1, TDP43 and GFAP datasets. This involved the following steps: handling missing values, normalisation, assessing constant features, and multicollinearity. The complete data preprocessing pipeline is depicted in Appendix Figure 2.

Polygon-based ROI annotations generated by QuPath’s superpixel segmentation algorithm^21^ were excluded because they represent segmentation metadata rather than quantitative pathological measurements and therefore do not constitute informative predictive features.

Highly correlated feature pairs were identified using pairwise Pearson correlation coefficients, with *|r| ≥* 0.95 considered indicative of strong collinearity. For the feature-reduction experiment, one feature from each highly correlated pair was removed using a deterministic pair-order rule, where the second feature in each identified pair was excluded. This procedure was applied independently to each biomarker-specific dataset. Model performance was then compared before and after removal of the selected collinear features.

Data normalisation is essential to ensure that feature values are on a common scale, enabling fair comparisons across the dataset by creating a standard range that prevents larger variables from dominating smaller ones^22^. MinMaxScaler was applied to normalise each feature within a fixed range of 0 to 1^22^. During fold-level model evaluation (3-fold or 5-group-CV), MinMax scaling was fitted using the training partition and subsequently applied to the corresponding validation partition. However, inspection of the original implementation indicated that for some hyperparameter-search procedures, scaling was performed prior to cross-validation. This represents a limitation of the current analytical pipeline and is considered when interpreting tuned-model performance.

Samples lacking either a class label or ROI annotation (n = 11) were excluded. Given the small number of incomplete observations, no imputation was performed. To evaluate the influence of low-information variables on model performance, constant features were identified prior to model development. Columns were considered constant if all values were zero or if the column contained only one unique value across the dataset. The union of these columns was removed to generate the constant-feature-reduced dataset. A comparative analysis was then performed using datasets before and after the exclusion of these features.

### 4.3 Machine Learning Pipeline

#### 4.3.1 Data Division

Two data division strategies were implemented. For benchmarking against the original study^11^, conventional stratified 3-fold cross-validation (3-CV) was implemented using shuffled folds with a fixed random seed (`random_state`=42). Stratification preserved the relative distribution of case and control observations across folds. However, observations were not grouped by individual, allowing samples from the same patient to occur in both training and validation folds, failing to account for intra-patient correlations and thereby introducing data leakage into the ML pipeline.

To improve robustness, a GroupKFold CV^23^ with 5 splits was also implemented, ensuring that samples from the same individual were kept together in either the training or test set, and dividing the group of 20 individuals into 5 non-overlapping folds. Note that 3-CV may lead to lower variability in model performance than 6-CV because fewer splits mean more individuals per split, making the results more homogeneous.

#### 4.3.2 Machine Learning models

To optimise the ALS identification process, an ML framework was implemented, as depicted in Figure 3. Due to the small sample size and high-dimensional nature of the data, different types of models were chosen based on their known strengths under such conditions. The ML models implemented are as follows:

- A random forest was used as a benchmark model, as it was also used in^11^, allowing direct performance comparison. Its ensemble nature, robustness to noise, and inherent feature selection capability make it ideal for high-dimensional biomedical data. Additionally, it allowed for feature selection by identifying the most important features.
- Support Vector Machines (SVMs) are widely recognised for their strong ability to handle binary classification tasks and their remarkable generalisation capability across diverse datasets^24^. This allows the model to create flexible decision boundaries that work well on complex datasets.
- Logistic regression, due to its simplicity, interpretability, and ability to generalise well with minimal risk of overfitting, makes it a suitable choice in this context^25^. Two solvers, liblinear and saga, were used because they support L1 and L2 regularisation penalties, which help prevent overfitting by reducing the importance of features that do not add much useful information^26^.
- XGBoost is a gradient boosting framework widely used for structured and tabular data. Its tree-based approach is well suited to high-dimensional datasets and can effectively model complex, non-linear relationships between features^27^.
- Artificial Neural Networks (ANNs) can model complex, non-linear relationships between features and class labels by learning hierarchical feature representations directly from the data. They jointly perform feature extraction and classification in an end-to-end manner, with the network parameters optimised through gradient-based learning using an appropriate loss function for the prediction task.
- An ensemble learning model using stacking was implemented to combine the strengths of all base classifiers. The predicted probabilities from the selected models were then used as inputs to a meta-classifier. A meta-classifier, in this case, logistic regression, provides an interpretable and efficient way to combine the base predictions^28^. It learns optimal weights for each base model’s output and produces probabilistic predictions that aid threshold tuning. Additionally, it is robust and less prone to overfitting at the meta-layer^28^.

#### 4.3.3 Hyperparameter Tuning

The hyperparameter tuning process was applied to all ML models using two approaches, grid and random search. Hyperparameter optimisation was performed using 3-fold stratified CV. For the random forest, RandomizedSearchCV evaluated 100 randomly selected hyperparameter combinations from the predefined search space, with accuracy used as the optimisation criterion. The hyperparameter configuration achieving the highest mean cross-validation accuracy was selected and subsequently used for model evaluation. For SVM, logistic regression, XGBoost, ANN, and the ensemble models, a grid search evaluated every possible combination of these values (Table 8).

**Table 8.** Hyperparameters tuned for each machine learning model, along with the corresponding search spaces explored during hyperparameter optimisation.

| Model | Parameter | Range of values |
| --- | --- | --- |
| Random forest | n_estimators<br>min_samples_split<br>min_samples_leaf<br>max_features<br>max_depth<br>bootstrap | [50, 100, 200]<br>[2, 5, 10]<br>[1, 2, 4]<br>[sqrt, log2, 0.5, None]<br>[None, 10, 20, 30]<br>[True, False] |
| SVM | C<br>gamma<br>kernel | [0.1, 1, 10, 100]<br>[scale, auto, 0.01]<br>[linear, rbf] |
| Logistic Regression | C | [0.001, 0.01, 0.1, 1, 10, 100] |
| XGBoost | penalty<br>solver<br>learning_rate<br>max_depth<br>n_estimators | [11, 12]<br>[liblinear]<br>[0.01, 0.1, 0.3]<br>[3, 5, 7]<br>[50, 100, 200] |
| ANN | batch_size<br>model_activation<br>model_layers<br>model_learning_rate<br>model_neurons | [16, 32]<br>[ReLU, tanh]<br>[1, 2]<br>[0.001, 0.01]<br>[64, 128] |
| Ensemble | final_estimator_C | [0.1, 1.0, 10] |

Abbreviations: ANN - Artificial Neural Network, SVM - Support Vector Machine ReLU - Rectified Linear Unit, RBF - Radial Basis Function, C - regularisation parameter.

Hyperparameter optimisation and performance estimation were not implemented using nested cross-validation. Consequently, the same dataset contributed to both model selection and subsequent performance estimation, which may result in optimistic estimates of tuned-model performance. The optimised hyperparameter values for each machine learning model-biomarker combination are reported in Table 9. Note that, for benchmarking purposes, hyperparameter tuning was performed in this study, whereas it was not applied in the previous study^11^. Furthermore, the previous study did not report the hyperparameter values used to train their models.

**Table 9.**
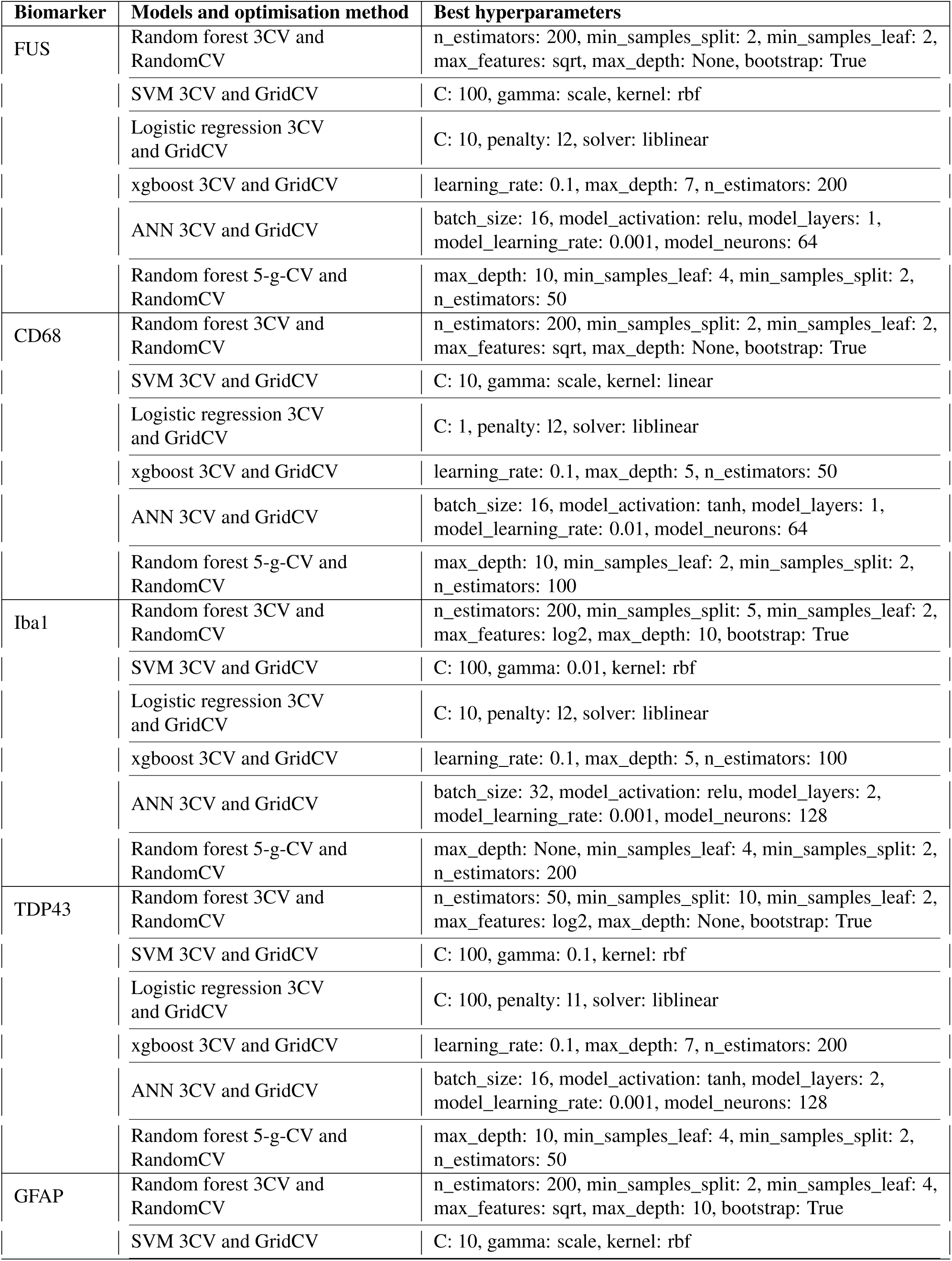
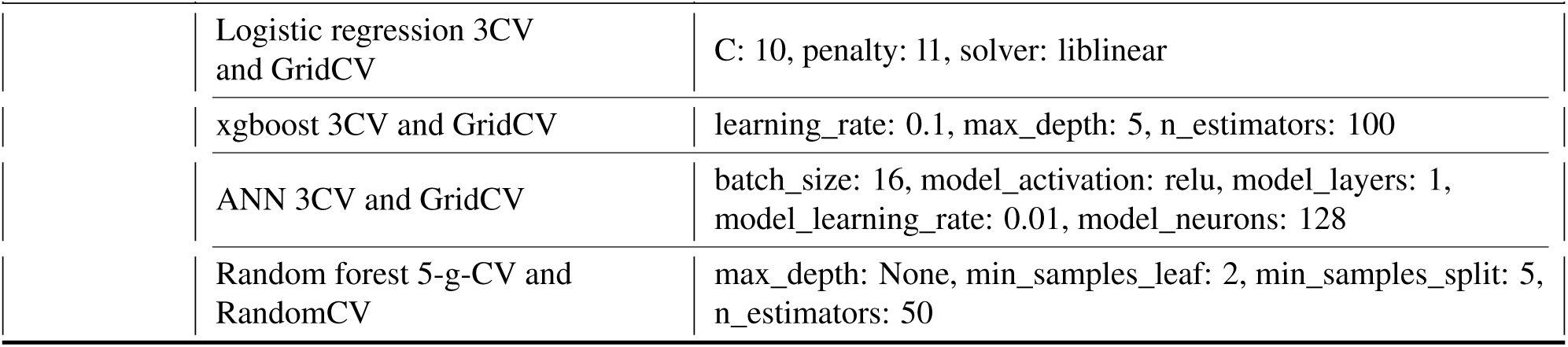
Optimised hyperparameters for different ML models across the five biomarkers.

| **Biomarker** | **Models and optimisation method** | **Best hyperparameters** |
| --- | --- | --- |
| FUS | Random forest 3CV and RandomCV | n_estimators: 200, min_samples_split: 2, min_samples_leaf: 2, max_features: sqrt, max_depth: None, bootstrap: True |
|  | SVM 3CV and GridCV | C: 100, gamma: scale, kernel: rbf |
|  | Logistic regression 3CV and GridCV | C: 10, penalty: l2, solver: liblinear |
|  | xgboost 3CV and GridCV | learning_rate: 0.1, max_depth: 7, n_estimators: 200 |
|  | ANN 3CV and GridCV | batch_size: 16, model_activation: relu, model_layers: 1, model_learning_rate: 0.001, model_neurons: 64 |
|  | Random forest 5-g-CV and RandomCV | max_depth: 10, min_samples_leaf: 4, min_samples_split: 2, n_estimators: 50 |
| CD68 | Random forest 3CV and RandomCV | n_estimators: 200, min_samples_split: 2, min_samples_leaf: 2, max_features: sqrt, max_depth: None, bootstrap: True |
|  | SVM 3CV and GridCV | C: 10, gamma: scale, kernel: linear |
|  | Logistic regression 3CV and GridCV | C: 1, penalty: l2, solver: liblinear |
|  | xgboost 3CV and GridCV | learning_rate: 0.1, max_depth: 5, n_estimators: 50 |
|  | ANN 3CV and GridCV | batch_size: 16, model_activation: tanh, model_layers: 1, model_learning_rate: 0.01, model_neurons: 64 |
|  | Random forest 5-g-CV and RandomCV | max_depth: 10, min_samples_leaf: 2, min_samples_split: 2, n_estimators: 100 |
| Iba1 | Random forest 3CV and RandomCV | n_estimators: 200, min_samples_split: 5, min_samples_leaf: 2, max_features: log2, max_depth: 10, bootstrap: True |
|  | SVM 3CV and GridCV | C: 100, gamma: 0.01, kernel: rbf |
|  | Logistic regression 3CV and GridCV | C: 10, penalty: l2, solver: liblinear |
|  | xgboost 3CV and GridCV | learning_rate: 0.1, max_depth: 5, n_estimators: 100 |
|  | ANN 3CV and GridCV | batch_size: 32, model_activation: relu, model_layers: 2, model_learning_rate: 0.001, model_neurons: 128 |
|  | Random forest 5-g-CV and RandomCV | max_depth: None, min_samples_leaf: 4, min_samples_split: 2, n_estimators: 200 |
| TDP43 | Random forest 3CV and RandomCV | n_estimators: 50, min_samples_split: 10, min_samples_leaf: 2, max_features: log2, max_depth: None, bootstrap: True |
|  | SVM 3CV and GridCV | C: 100, gamma: 0.1, kernel: rbf |
|  | Logistic regression 3CV and GridCV | C: 100, penalty: l1, solver: liblinear |
|  | xgboost 3CV and GridCV | learning_rate: 0.1, max_depth: 7, n_estimators: 200 |
|  | ANN 3CV and GridCV | batch_size: 16, model_activation: tanh, model_layers: 2, model_learning_rate: 0.001, model_neurons: 128 |
|  | Random forest 5-g-CV and RandomCV | max_depth: 10, min_samples_leaf: 4, min_samples_split: 2, n_estimators: 50 |
| GFAP | Random forest 3CV and RandomCV | n_estimators: 200, min_samples_split: 2, min_samples_leaf: 4, max_features: sqrt, max_depth: 10, bootstrap: True |
|  | SVM 3CV and GridCV | C: 10, gamma: scale, kernel: rbf |

| Biomarker | Models and optimisation method | Best hyperparameters |
| --- | --- | --- |
|  | Logistic regression 3CV and GridCV | C: 10, penalty: 11, solver: liblinear |
|  | xgboost 3CV and GridCV | learning_rate: 0.1, max_depth: 5, n_estimators: 100 |
|  | ANN 3CV and GridCV | batch_size: 16, model_activation: relu, model_layers: 1, model_learning_rate: 0.01, model_neurons: 128 |
|  | Random forest 5-g-CV and RandomCV | max_depth: None, min_samples_leaf: 2, min_samples_split: 5, n_estimators: 50 |

##### Model configuration and training

In addition to the hyperparameters included in the optimisation procedure, model-specific parameters were fixed during training to ensure reproducibility. A random seed of 42 was used where supported. For the baseline random forest model, 100 trees were used. The baseline SVM used a linear kernel with *C* = 1.0, while probability estimation was enabled for SVM when used as a base learner in the stacking ensemble. Logistic regression used the corresponding scikit-learn implementation, with parameters not included in the optimisation procedure left at their default values unless otherwise specified. XGBoost used log-loss as the evaluation metric.

The ANN used the Adam optimiser and binary cross-entropy loss, with a single sigmoid-activated output unit for binary classification. A dropout rate of 0.2 was applied to the hidden layers. The number of hidden layers, number of neurons, hidden-layer activation function, learning rate, and batch size were included in the hyperparameter search, whereas the number of training epochs was fixed rather than optimised. During hyperparameter optimisation, ANN models were trained for a maximum of 100 epochs. The selected ANN configuration was subsequently evaluated with a maximum of 50 epochs.

The stacking ensemble combined random forest, SVM, logistic regression, XGBoost and ANN base learners, with logistic regression used as the meta-classifier. The SVM base learner was configured to generate probability estimates. Two-fold stratified cross-validation was used internally by the stacking procedure to generate predictions from the base learners. The regularisation parameter *C* of the logistic-regression meta-classifier was optimised over the values [0.1, 1.0, 10]. The ANN base learner within the ensemble was trained for a maximum of 30 epochs using a batch size of 32 and a validation split of 0.1, with early stopping using a patience of five epochs and restoration of the best model weights. Model parameters not explicitly optimised or specified above were left at their respective library default values.

### 4.4 Model Training and Performance Evaluation

To assess model performance, the same evaluation metrics as in^11^ were used. Each model was evaluated using 3-CV, while 5-g-CV was implemented for the random forest model and ensemble models. The objective was to evaluate the performance of the models after removing data leakage to obtain more realistic results. However, a limitation of the current version is that the models within the ensemble also suffer from data leakage. When these models are ensembled, the problem is inherited, as the individual models retain the leakage issue during stacking. The mean and SD of accuracy, sensitivity, and specificity across folds were reported to summarise model performance and to ensure stable benchmarking against the baseline methods^11^. Note that, for comparison purposes, we manually calculated the accuracy of the results reported by Rifai *et al.*, as this metric was not provided in the original study. This was possible because the dataset contained an equal number of samples in each class.

Checking for statistical significance is a standard procedure in the medical domain to ensure the reliability of model performance. To enable a direct comparison with the previous study, the same statistical validation strategy was adopted. This analysis aimed to confirm that the observed differences in brain biomarkers were associated with disease status rather than random variation.

The Shapiro–Wilk test was first used to assess the normality of each performance metric and determine the appropriate statistical test. When *p >* 0.05, the data were considered normally distributed, and a Chi-squared test was subsequently applied. A Chi-squared test result of *p <* 0.05 indicated that the classification performance was significantly better than chance. When the Shapiro–Wilk test indicated that the normality assumption was violated (*p ≤* 0.05), the Wilcoxon signed-rank test was applied instead. For each performance metric, the Wilcoxon test evaluated whether the median performance differed significantly from the chance-level value of 0.5, with statistical significance defined as *p <* 0.05.

#### Feature importance analysis

Feature importance was evaluated using the random forest models trained under the 5-grouped cross-validation (5-g-CV) framework. For each fold, feature importance scores were extracted from the fitted random forest using the mean decrease in Gini impurity. The resulting importance values were averaged across the five folds to obtain a mean importance score for each feature. Variability in feature importance across folds was not reported in the original analysis, and the evaluation was based on a single grouped cross-validation partition. Features were then ranked independently for each biomarker-specific model (FUS, CD68, Iba1, TDP43 and GFAP), and the ten features with the highest mean importance scores were retained for interpretation.

Mean decrease in Gini impurity is model-dependent and may be influenced by feature variability and correlation between predictors. In the presence of highly correlated features, importance can be distributed across variables carrying similar information, which may reduce or destabilise the apparent importance of individual features. Feature importance values were therefore interpreted as relative indicators of contribution within each random forest model rather than as direct measures of biological relevance.

This approach differs from the feature analysis performed by Rifai *et al.*^11^. In addition to evaluating the biomarkers individually, Rifai *et al.* constructed a random forest model incorporating features across the biomarker panel to investigate their contribution to disease-status prediction. Feature contribution was assessed using a leave-one-feature-out approach, in which individual features were removed, and the resulting change in predictive performance was evaluated. The highest-ranking features were subsequently examined according to six feature categories: cell morphology, vessel-related features, FUS cell staining, superpixel spatial distribution, superpixel staining and superpixel morphology. In contrast, the present study calculates Gini-based feature importance independently within each biomarker-specific random forest model. Consequently, the two approaches address different questions, and their feature-importance values are not directly comparable: the analysis of Rifai *et al.* examines feature contribution across the combined pathological feature space, whereas the present analysis identifies the features contributing most strongly to classification within each individual biomarker.

## Conclusion and Future Work

This study reconstructed five structured, ML-ready datasets from the digital pathology measurements reported by Rifai *et al.*^11^ and extended their random forest analysis to a broader range of machine-learning models. The results demonstrate that classification performance is both biomarker- and model-dependent, with no single algorithm consistently achieving the highest sensitivity, specificity and accuracy across all biomarkers. Iba1 showed the strongest overall predictive performance and remained the strongest random forest biomarker when patient-level separation was introduced, supporting the predictive relevance of this marker observed in the baseline study. The biomarker-specific feature-importance analysis further indicated that classification relies on combinations of pathological measurements rather than a single dominant feature, although interpretation of individual feature rankings is limited by multicollinearity.

A central finding was the systematic reduction in random forest performance when conventional 3-fold cross-validation was replaced by patient-grouped 5-fold cross-validation. This demonstrates that validation design can substantially influence apparent model performance in datasets containing multiple observations from the same individuals. However, patient grouping alone does not provide a completely independent estimate of generalisation: hyperparameter optimisation was not fully nested within the patient-grouped evaluation, and some data-dependent preprocessing was performed outside the corresponding cross-validation folds. Furthermore, the other classifiers were not evaluated under the same patient-grouped framework. Consequently, the current results do not establish which algorithm generalises best to previously unseen patients.

Future work should therefore prioritise repeated, fully nested patient-grouped cross-validation across all classifiers, ensuring that preprocessing, hyperparameter optimisation and model training are restricted to the corresponding training patients. This framework would enable more reliable estimation of performance variability, statistical comparison between models, and assessment of the stability of feature rankings. Further extensions should investigate robust treatment of near-constant and correlated features, improved ANN training with appropriately implemented early stopping, and a fully patient-isolated stacking ensemble. Validation in larger independent cohorts will ultimately be required to establish the generalisability of the identified pathological signals. The available histopathological images also provide an opportunity for subsequent image-based and multimodal analyses combining learned image representations with the quantitative pathological features investigated here.

## Data Ethics

Ethical approvals for data and tissue collection were obtained from the East of Scotland Research Ethics Service (16/ES/0084), in accordance with the Human Tissue (Scotland) Act (2006). The use of post-mortem samples was reviewed and approved by the Edinburgh Brain Bank Ethics Committee and the Academic and Clinical Central Office for Research and Development (AMREC). Clinical data were collected through the Scottish Motor Neuron Disease Register (SMNDR) and the Care Audit Research and Evaluation for Motor Neuron Disease (CARE-MND) platform, with ethical approval from the Scotland A Research Ethics Committee (10/MRE00/78 and 15/SS/0216). All patients provided informed consent for the use of their data for research while they were alive, and clinical and cognitive data such as ECAS and ALSFRS were collected^11^. The owners of the data granted permission to use this dataset for secondary processing.

## Data Availability

This study is a secondary analysis of previously published, ethically approved post-mortem brain tissue data obtained from publicly available figshare repositories linked in the original publication (cite: Rifai, O.M., Longden, J., O'Shaughnessy, J., Sewell, M.D., Pate, J., McDade, K., Daniels, M.J., Abrahams, S., Chandran, S., McColl, B.W. and Sibley, C.R., 2022. Random forest modelling demonstrates microglial and protein misfolding features to be key phenotypic markers in C9orf72‐ALS. The Journal of pathology, 258(4), pp.366-381.).
The SD numbers of cases from the Edinburgh Brain Bank included in the study are available upon request.

https://figshare.com/projects/Random_forest_modelling_and_neuropathological_features_of_a_large_cohort_of_C9orf72-ALS_all_raw_data_/128222

https://doi.org/10.6084/m9.figshare.17145902.v1

https://pathsocjournals.onlinelibrary.wiley.com/action/downloadSupplement?doi=10.1002%2Fpath.6008&file=path6008-sup-0001-SuppMatMeth%2CFiguresS1-S2%2CTableS1-S4.docx

https://figshare.com/articles/dataset/Random_forest_results/17145905

## Appendix

**Table 10.** Computation of the pathological features reconstructed from the original QuPath-derived measurements.

| Feature/s | Biomarker | Files involved (.csv) | Calculation |
| --- | --- | --- | --- |
| Area (px<sup>2</sup>) of vessel (VA) | FUS, CD68, Iba1, TDP43, GFAP | VA_annotations | Per unique image (VA) (case/control) – mean of area (px<sup>2</sup>) |
| Average area (px<sup>2</sup>) of class - 1+, 2+, 3+ and negative superpixels | FUS, CD68, Iba1, TDP43, GFAP | NVA_detections, VA_detections_normed | Per-class mean of area (px<sup>2</sup>) |
| Average circularity of class - 1+, 5+, 3+ and negative superpixels | FUS, CD68, Iba1, TDP43, GFAP | NVA_detections, VA_detections_normed | Per-class mean of circularity |
| Average distance (px) of class 1+, 2+, 3+, negative superpixels to class 1+, 2+, 3+, negative superpixels | FUS, CD68, Iba1, TDP43, GFAP | NVA_detections, VA_detections_normed | Per-class mean of distance (px) from class (1+, 2+, 3+, negative) to class (1+, 2+, 3+, negative) |
| Average glial cell area | FUS | FUS_NVA_glia, FUS_VA_glia | Per unique image (case/control) – mean of glial cell area |
| Average glial cell circularity | FUS | FUS_NVA_glia, FUS_VA_glia | Per unique image (case/control) – mean of glial cell circularity |
| Average glial cell DAB intensity (max, min, mean, and standard deviation) | FUS | FUS_NVA_glia, FUS_VA_glia | Per unique image (case/control) – mean of glial cell DAB intensity (max, min, mean, and standard deviation) |
| Average glial cell eccentricity | FUS | FUS_NVA_glia, FUS_VA_glia | Per unique image (case/control) – mean of glial cell eccentricity |
| Average glial cell haematoxylin intensity (max, min, mean, standard deviation) | FUS | FUS_NVA_glia, FUS_VA_glia | Per unique image (case/control) – mean of glial cell haematoxylin intensity (max, min, mean, standard deviation) |
| Average glial cell max/min calliper | FUS | FUS_NVA_glia, FUS_VA_glia | Per unique image (case/control) – mean of glial cell max/min calliper |
| Average glial cell perimeter | FUS | FUS_NVA_glia, FUS_VA_glia | Per unique image (case/control) – mean of glial cell perimeter |
| Average glial cytoplasmic DAB intensity (max/ min/ mean/ standard deviation) | FUS | FUS_NVA_glia, FUS_VA_glia | Per unique image (case/control) – mean of glial cytoplasmic DAB intensity (max/ min/ mean/ standard deviation) |
| Average glial cytoplasmic haematoxylin intensity (max/ min/ range/ standard deviation) | FUS | FUS_NVA_glia, FUS_VA_glia | Per unique image (case/control) – mean of glial cytoplasmic haematoxylin intensity (max/ min/ range/ standard deviation) |
| Average glial nuclear area | FUS | FUS_NVA_glia, FUS_VA_glia | Per unique image (case/control) – mean of glial nuclear area |
| Average glial nuclear circularity | FUS | FUS_NVA_glia, FUS_VA_glia | Per unique image (case/control) – mean of glial nuclear circularity |
| Average glial nuclear DAB intensity (max/ min/ mean/ range/ standard deviation/ sum) | FUS | FUS_NVA_glia, FUS_VA_glia | Per unique image (case/control) – mean of glial nuclear DAB intensity (max/ min/ mean/ range/ standard deviation/ sum) |
| Average glial nuclear eccentricity | FUS | FUS_NVA_glia, FUS_VA_glia, | Per unique image (case/control) – mean of glial nuclear eccentricity |
| Average glial nuclear haematoxylin intensity (max/ min/ mean/ range/ standard deviation/ sum) | FUS | FUS_NVA_glia, FUS_VA_glia, | Per unique image (case/control) – mean of glial nuclear haematoxylin intensity (max/ min/ mean/ range/ standard deviation/ sum) |
| Average glial nuclear max/min calliper | FUS | FUS_NVA_glia, FUS_VA_glia, | Per unique image (case/control) – mean of glial nuclear max/min calliper |
| Average glial nuclear perimeter | FUS | FUS_NVA_glia, FUS_VA_glia, | Per unique image (case/control) – mean of glial nuclear perimeter |
| Average glial nuclear/cytoplasmic DAB intensity ratio | FUS | FUS_NVA_glia, FUS_VA_glia, | Per unique image (case/control) – mean of glial nuclear/cytoplasmic DAB intensity ratio |
| Average glial nucleus/cell area ratio | FUS | FUS_NVA_glia, FUS_VA_glia, | Per unique image (case/control) – mean of glial nucleus/cell area ratio |
| Average length (px) of class -1+, 2+, 3+ and negative superpixels | FUS, CD68, Iba1, TDP43, GFAP | NVA_detections, VA_detections_normed | Per-class average of length |
| Average max/min diameter (px) of class 1+, 2+, 3+ and negative superpixels | FUS, CD68, Iba1, TDP43, GFAP | NVA_detections, VA_detections_normed | Per-class average of max/min diameter |
| Average neuronal cell area | FUS | NVA_neurons, VA_neurons | Per unique image (case/control) – mean of neuronal cell area |
| Average neuronal cell circularity | FUS | NVA_neurons, VA_neurons | Per unique image (case/control) – mean of neuronal cell circularity |
| Average neuronal cell DAB intensity(max/ min/ mean /standard deviation) | FUS | NVA_neurons, VA_neurons | Per unique image (case/control) – mean of neuronal cell DAB intensity (max/ min/ mean /standard deviation) |
| Average neuronal cell eccentricity | FUS | NVA_neurons, VA_neurons | Per unique image (case/control) –mean of neuronal cell eccentricity |
| Average neuronal cell haematoxylin intensity (max / min/ mean/ standard deviation) | FUS | NVA_neurons, VA_neurons | Per unique image (case/control) – mean of neuronal cell haematoxylin intensity (max / min/ mean/ standard deviation) |
| Average neuronal cell eccentricity | FUS | NVA_neurons, VA_neurons | Per unique image (case/control) – mean of neuronal cell eccentricity |
| Average neuronal cell haematoxylin intensity (max / min/ mean/ standard deviation) | FUS | NVA_neurons, VA_neurons | Per unique image (case/control) – mean of neuronal cell haematoxylin intensity (max / min/ mean/ standard deviation) |
| Average neuronal cell max/ min calliper | FUS | NVA_neurons, VA_neurons | Per unique image (case/control) – mean of neuronal cell max/ min calliper |
| Average neuronal cell perimeter | FUS | NVA_neurons,<br>VA_neurons | Per unique image (case/control) – mean of neuronal cell perimeter |
| Average neuronal cytoplasmic DAB intensity (max/ min /mean/ standard deviation) | FUS | NVA_neurons,<br>VA_neurons | Per unique image (case/control) – mean of neuronal cytoplasmic DAB intensity (max/ min /mean/ standard deviation) |
| Average neuronal cytoplasmic haematoxylin intensity (max/ min/ mean/ standard deviation) | FUS | NVA_neurons,<br>VA_neurons | Per unique image (case/control) – mean of neuronal cytoplasmic haematoxylin intensity (max/ min/ mean/ standard deviation) |
| Average neuronal haematoxylin intensity (mean) | FUS | NVA_neurons,<br>VA_neurons | Per unique image (case/control) – mean of neuronal haematoxylin intensity (mean) |
| Average neuronal nuclear area | FUS | NVA_neurons,<br>VA_neurons | Per unique image (case/control) – mean of neuronal nuclear area |
| Average neuronal nuclear cell area ratio | FUS | NVA_neurons,<br>VA_neurons | Per unique image (case/control) – mean of neuronal nuclear cell area ratio |
| Average neuronal nuclear circularity | FUS | NVA_neurons,<br>VA_neurons | Per unique image (case/control) – mean of neuronal nuclear circularity |
| Average neuronal nuclear DAB intensity (max/ min/ mean/ range/ standard deviation/ sum) | FUS | NVA_neurons,<br>VA_neurons | Per unique image (case/control) – mean of neuronal nuclear DAB intensity (max/ min/ mean/ range/ standard deviation/ sum) |
| Average neuronal nuclear eccentricity | FUS | NVA_neurons,<br>VA_neurons | Per unique image (case/control) – mean of neuronal nuclear eccentricity |
| Average neuronal nuclear haematoxylin intensity (max/ min/ range/ standard deviation/ sum) | FUS | NVA_neurons,<br>VA_neurons | Per unique image (case/control) – mean of neuronal nuclear haematoxylin intensity (max/ min/ range/ standard deviation/ sum) |
| Average neuronal nuclear max/min caliper | FUS | NVA_neurons,<br>VA_neurons | Per unique image (case/control) – mean of neuronal nuclear max/min caliper |
| Average neuronal nuclear perimeter | FUS | NVA_neurons,<br>VA_neurons | Per unique image (case/control) – mean of neuronal nuclear perimeter |
| Average neuronal nuclear/ cytoplasmic DAB intensity ratio | FUS | NVA_neurons,<br>VA_neurons | Per unique image (case/control) – mean of neuronal nuclear/ cytoplasmic DAB intensity ratio |
| Average normalised distance (px) of class 1+, 2+, 3+ and negative superpixels to vessel (VA) | FUS, CD68, Iba1, TDP43, GFAP | NVA_detections_norm | Per-class average of normalised distance |
| Average solidity of class 1+, 2+, 3+ and negative superpixels | FUS, CD68, Iba1, TDP43, GFAP | NVA_detections, VA_detections_norm | Per-class average of solidity |
| Image Allred intensity | FUS, CD68, Iba1, TDP43, GFAP | NVA_annotations, VA_annotations | Direct feature retrieval |
| Image Allred proportion | FUS, CD68, Iba1, TDP43, GFAP | NVA_annotations, VA_annotations | Direct feature retrieval |
| Image Allred score | FUS, CD68, Iba1, TDP43, GFAP | NVA_annotations, VA_annotations | Direct feature retrieval |
| Image H-score | FUS, CD68, Iba1, TDP43, GFAP | NVA_annotations, VA_annotations | Direct feature retrieval |
| Image area excluding vessel (px<sup>2</sup>) | FUS, CD68, Iba1, TDP43, GFAP | NVA_annotations, VA_annotations, VA_polygons_vessel | $Area_{px^2} - Vessel_{area}$ |
| Percentage of super-pixels that are class 1+, 2+, 3+ and negative (VA) | FUS, CD68, Iba1, TDP43, GFAP | NVA_detections_norm | (Count of class per image (VA)) / (Total counts for that image (VA)) × 100 |
| Percentage of super-pixels that are class 1+, 2+, 3+ and negative | FUS, CD68, Iba1, TDP43, GFAP | NVA_detections, VA_detections_norm | (Count of class per image) / (Total counts for that image) × 100 |
| Perimeter (px) of vessel (VA) | FUS, CD68, Iba1, TDP43, GFAP | VA_polygons_vessels | Direct feature retrieval |
| Vessel Allred intensity (VA) | FUS, CD68, Iba1, TDP43, GFAP | VA_polygons_vessels | Direct feature retrieval |
| Vessel Allred proportion (VA) | FUS, CD68, Iba1, TDP43, GFAP | VA_polygons_vessels | Direct feature retrieval |
| Vessel Allred score (VA) | FUS, CD68, Iba1, TDP43, GFAP | VA_polygons_vessels | Direct feature retrieval |
| Vessel area (VA) | FUS, CD68, Iba1, TDP43, GFAP | VA_polygons_vessels | Direct feature retrieval |
| Vessel H-score (VA) | FUS, CD68, Iba1, TDP43, GFAP | VA_polygons_vessels | Direct feature retrieval |

**Table 11.** Comparison of random forest classification performance reported by Rifai *et al.*^11^ with the tuned random forest models evaluated in the present study using conventional 3-fold cross-validation (3-CV) and patient-grouped 5-fold cross-validation (5-g-CV). Accuracy was not reported by Rifai *et al.* and was calculated from the reported sensitivity and specificity values, given the balanced number of observations in the two classes. Results from the present study are reported as mean *±* variability as provided by the current analysis. The different validation strategies should be considered when interpreting comparisons between studies.

| Biomarker | Study / validation | Sensitivity | Specificity | Accuracy |
| --- | --- | --- | --- | --- |
| FUS | Rifai *et al.* | 0.64 | 0.69 | 0.665 |
| | Current study, RF 3-CV | 0.73 ± 0.03 | 0.74 ± 0.03 | 0.73 ± 0.03 |
| | Current study, RF 5-g-CV | 0.592 ± 0.10 | 0.66 ± 0.13 | 0.624 ± 0.04 |
| CD68 | Rifai *et al.* | 0.67 | 0.60 | 0.635 |
| | Current study, RF 3-CV | 0.62 ± 0.01 | 0.76 ± 0.02 | 0.69 ± 0.01 |
| | Current study, RF 5-g-CV | 0.57 ± 0.07 | 0.69 ± 0.07 | 0.63 ± 0.07 |
| Iba1 | Rifai *et al.* | 0.78 | 0.84 | 0.81 |
| | Current study, RF 3-CV | 0.88 ± 0.03 | 0.83 ± 0.01 | 0.86 ± 0.01 |
| | Current study, RF 5-g-CV | 0.83 ± 0.06 | 0.76 ± 0.12 | 0.80 ± 0.06 |
| TDP43 | Rifai *et al.* | 0.51 | 0.64 | 0.575 |
| | Current study, RF 3-CV | 0.62 ± 0.03 | 0.69 ± 0.08 | 0.66 ± 0.04 |
| | Current study, RF 5-g-CV | 0.56 ± 0.11 | 0.60 ± 0.06 | 0.58 ± 0.05 |
| GFAP | Rifai *et al.* | 0.79 | 0.53 | 0.66 |
| | Current study, RF 3-CV | 0.70 ± 0.05 | 0.70 ± 0.03 | 0.70 ± 0.02 |
| | Current study, RF 5-g-CV | 0.60 ± 0.10 | 0.68 ± 0.09 | 0.64 ± 0.08 |

**Table 12.**
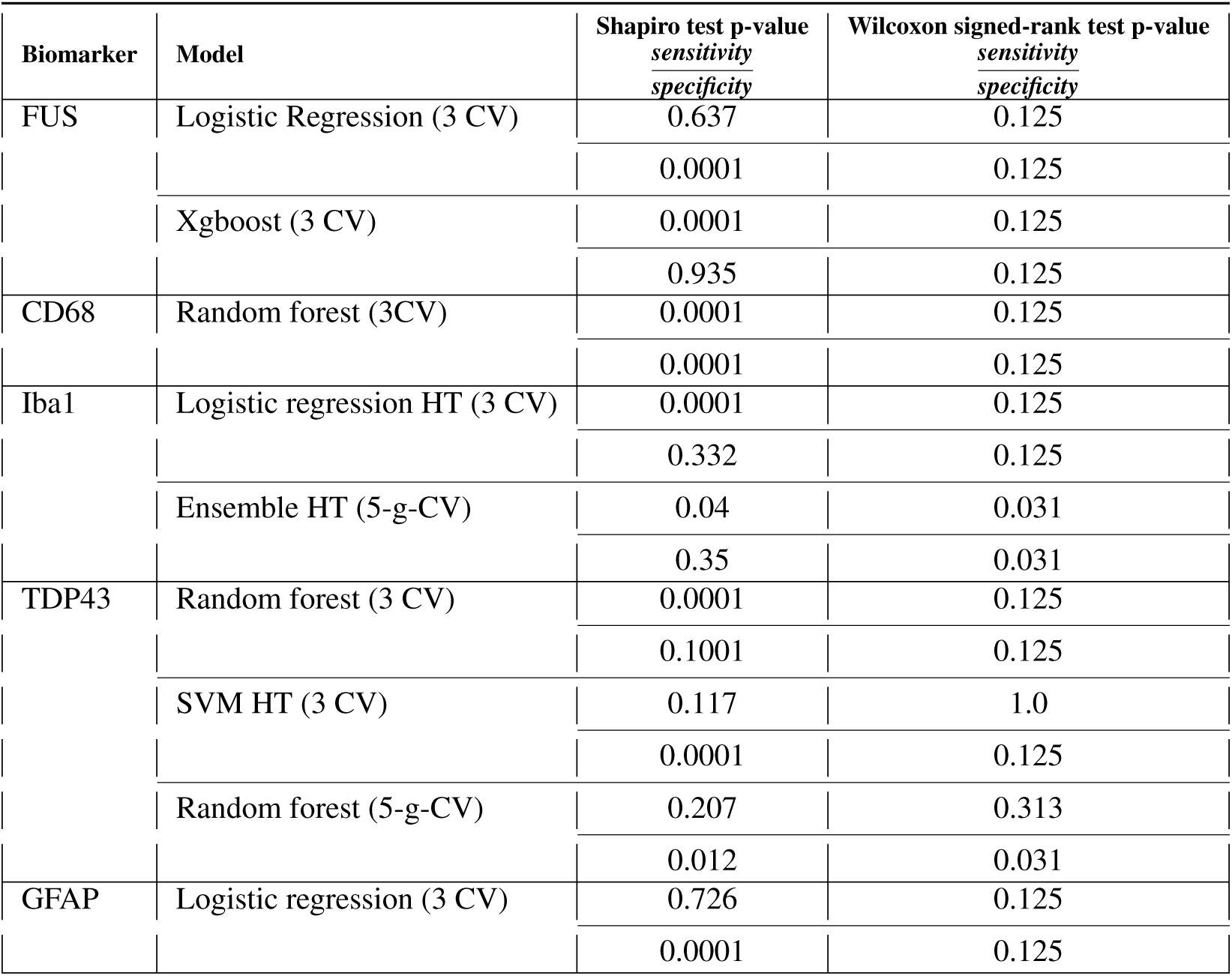
Shapiro test and Wilcoxon test p-values for the models when the Shapiro test failed.

**Table 13.** Model performance following the removal of constant features from the biomarker datasets.

| Biomarker | Model | Shapiro test p-values<br>$\frac{\text{sensitivity}}{\text{specificity}}$ | Sensitivity<br>($\mu \pm \sigma$) | Specificity<br>($\mu \pm \sigma$) | Chi<sup>2</sup> test<br>p-value | Accuracy<br>($\mu \pm \sigma$) |
| --- | --- | --- | --- | --- | --- | --- |
| FUS | RF<sup>1</sup> | $\frac{0.003}{0.007}$ | (0.72, ± 0.02) | (0.75, ± 0.04) | 0.0001 | (0.73, ± 0.03) |
| | RF<sup>2</sup> | $\frac{0.068}{0.029}$ | (0.6, ± 0.11) | (0.66, ± 0.12) | 0.0001 | (0.63, ± 0.04) |
| CD68 | RF<sup>1</sup> | $\frac{0.0036}{0.0004}$ | (0.64, ± 0.02) | (0.74, ± 0.01) | 0.0001 | (0.69, ± 0.01) |
| | RF<sup>2</sup> | $\frac{0.05}{0.005}$ | (0.58, ± 0.08) | (0.68, ± 0.08) | 0.0001 | (0.63, ± 0.07) |
| Iba1 | RF<sup>1</sup> | $\frac{0.0023}{0.0008}$ | (0.88, ± 0.04) | (0.83, ± 0.02) | 0.0001 | (0.85, ± 0.01) |
| | RF<sup>2</sup> | $\frac{0.0001}{0.005}$ | (0.83, ± 0.06) | (0.76, ± 0.11) | 0.0001 | (0.8, ± 0.06) |
| TDP43 | RF<sup>1</sup> | $\frac{0.0059}{0.0324}$ | (0.65, ± 0.02) | (0.62, ± 0.06) | 0.0001 | (0.63, ± 0.02) |
| | RF<sup>2</sup> | $\frac{0.113}{0.005}$ | (0.57, ± 0.1) | (0.63, ± 0.06) | 0.0001 | (0.6, ± 0.05) |
| GFAP | RF<sup>1</sup> | $\frac{0.035}{0.0028}$ | (0.72, ± 0.07) | (0.67, ± 0.02) | 0.0001 | (0.7, ± 0.03) |
| | RF<sup>2</sup> | $\frac{0.045}{0.019}$ | (0.61, ± 0.1) | (0.66, ± 0.1) | 0.0001 | (0.64, ± 0.09) |

$\mu$ - Mean, $\sigma$ - standard deviation, CV - Cross validation, ANN - Artificial Neural Network, \* - failed Shapiro test
<sup>1</sup>Model without hyperparameter tuning and 3 cross-validation
<sup>2</sup>Model with hyperparameter tuning and 3 cross-validation
<sup>3</sup>Model with 5-grouped cross-validation and hyperparameter tuning
<sup>4</sup>Model with 5-grouped cross-validation and without hyperparameter tuning

**Table 14.**
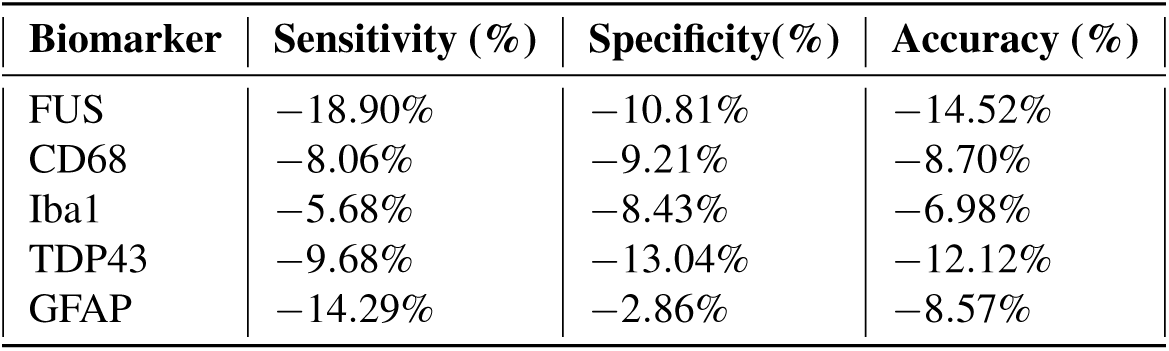
Percentage change in performance metrics of the patient-grouped 5-fold cross-validation (5-g-CV) random forest model compared with the conventional 3-fold cross-validation (3-CV) random forest model. Both models were evaluated with hyperparameter tuning. Negative values indicate a reduction in performance under patient-grouped cross-validation.

**Table 15.**
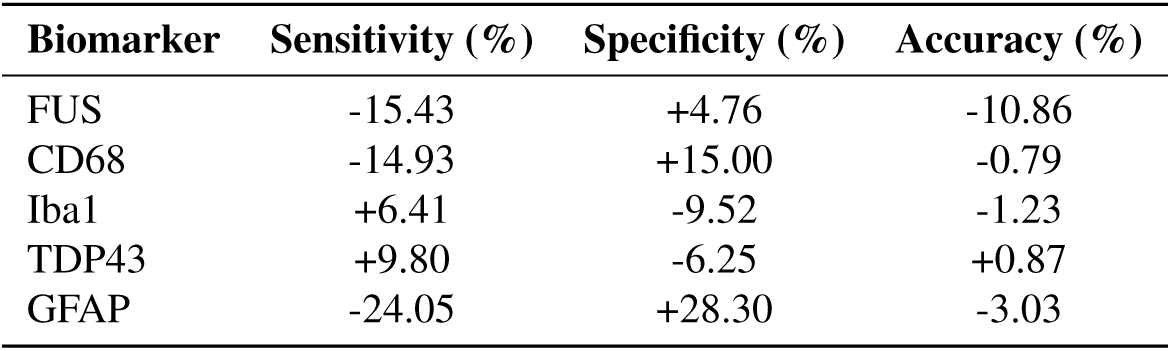
Comparison of the tuned 5-grouped cross-validation (5-g-CV) random forest model with the random forest benchmark reported by Rifai *et al.*^11^. Values represent the percentage change in performance relative to the benchmark. Positive values indicate improved performance, whereas negative values indicate reduced performance. Benchmark accuracy was calculated in the present study from the sensitivity and specificity values reported by Rifai *et al.*^11^, as accuracy was not reported in the original study.

| Biomarker | Sensitivity (%) | Specificity (%) | Accuracy (%) |
| --- | --- | --- | --- |
| FUS | -15.43 | +4.76 | -10.86 |
| CD68 | -14.93 | +15.00 | -0.79 |
| Iba1 | +6.41 | -9.52 | -1.23 |
| TDP43 | +9.80 | -6.25 | +0.87 |
| GFAP | -24.05 | +28.30 | -3.03 |

**Table 16.** Model performance after the removal of collinear columns from the biomarker datasets.

| Biomarker | Model | Shapiro test p-values<br>$\frac{\text{sensitivity}}{\text{specificity}}$ | Sensitivity<br>($\mu \pm \sigma$) | Specificity<br>($\mu \pm \sigma$) | Chi<sup>2</sup> test<br>p-value | Accuracy<br>($\mu \pm \sigma$) |
| --- | --- | --- | --- | --- | --- | --- |
| FUS | RF<sup>1</sup> | $\frac{0.0035}{0.0134}$ | (0.72, ± 0.03) | (0.71, ± 0.05) | 0.0001 | (0.71, ± 0.04) |
| | RF<sup>2</sup> | $\frac{0.023}{0.133}$ | (0.62, ± 0.08) | (0.63, ± 0.20) | 0.0001 | (0.62, ± 0.07) |
| CD68 | RF<sup>1</sup> | $\frac{0.0003}{0.0014}$ | (0.65, ± 0.01) | (0.74, ± 0.02) | 0.0001 | (0.70, ± 0.01) |
| | RF<sup>2</sup> | $\frac{0.05}{0.007}$ | (0.58, ± 0.08) | (0.65, ± 0.07) | 0.0001 | (0.62, ± 0.07) |
| Iba1 | RF<sup>1</sup> | $\frac{0.0028}{0.0005}$ | (0.84, ± 0.02) | (0.87, ± 0.04) | 0.0001 | (0.86, ± 0.02) |
| | RF<sup>2</sup> | $\frac{0.0001}{0.006}$ | (0.83, ± 0.05) | (0.76, ± 0.12) | 0.0001 | (0.80, ± 0.07) |
| TDP43 | RF<sup>1</sup> | $\frac{0.06}{0.02}$ | (0.58, ± 0.04) | (0.70, ± 0.02) | 0.0001 | (0.64, ± 0.03) |
| | RF<sup>2</sup> | $\frac{0.552}{0.005}$ | (0.5, ± 0.14) | (0.66, ± 0.07) | 0.0001 | (0.58, ± 0.05) |
| GFAP | RF<sup>1</sup> | $\frac{0.018}{0.0028}$ | (0.69, ± 0.05) | (0.72, ± 0.02) | 0.0001 | (0.70, ± 0.02) |
| | RF<sup>2</sup> | $\frac{0.045}{0.019}$ | (0.61, ± 0.10) | (0.66, ± 0.10) | 0.0001 | (0.64, ± 0.09) |

<sup>1</sup>Model with hyperparameter and 3 cross-validation
<sup>2</sup>Model with hyperparameter and 5-grouped cross-validation

**Table 17.**
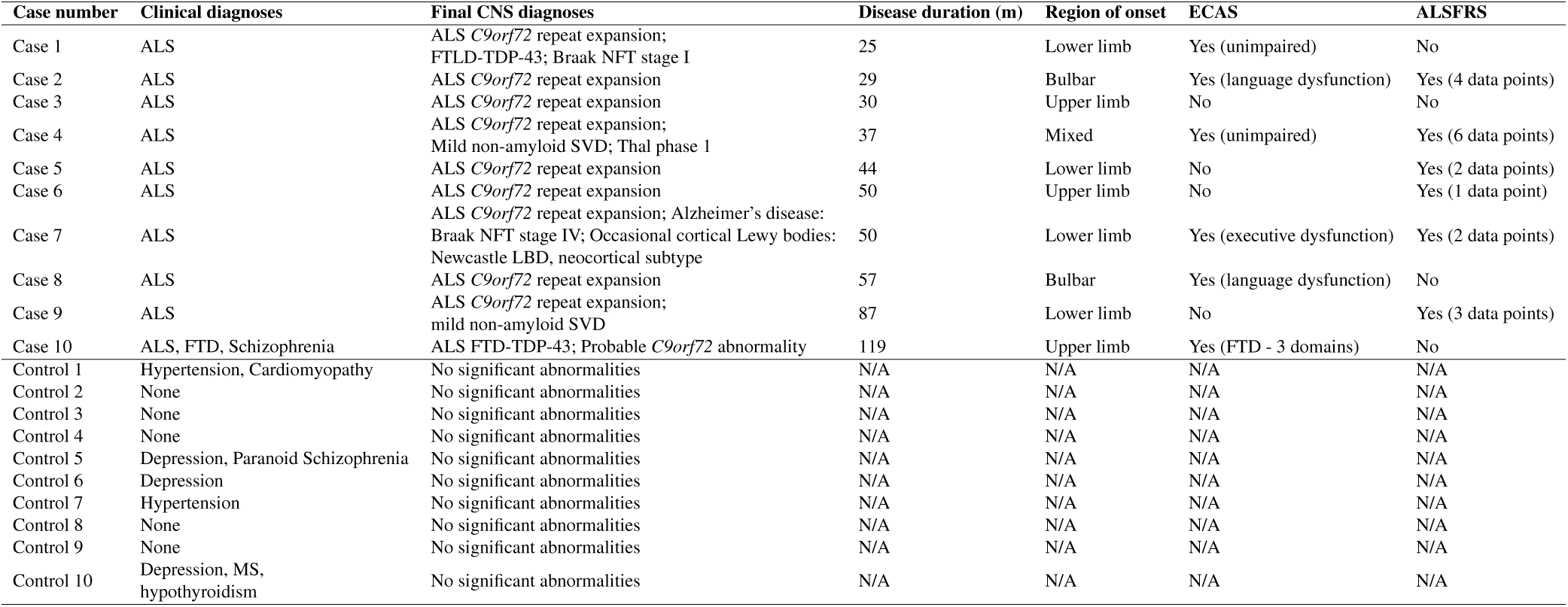
Clinical, neuropathological and disease characteristics of the C9-ALS and control cohort.

## Notes

### Competing Interest Statement

The authors have declared no competing interest.

### Author Declarations

The study used ONLY anonymised post-mortem brain tissue data obtained from openly available source located at: https://figshare.com/projects/Random_forest_modelling_and_neuropathological_features_of_a_large_cohort_of_C9orf72-ALS_all_raw_data_/128222. Data Ethics Ethical approvals for data and tissue collection were obtained from the East of Scotland Research Ethics Service (16/ES/0084), in accordance with the Human Tissue (Scotland) Act (2006). The use of post-mortem samples was reviewed and approved by the Edinburgh Brain Bank Ethics Committee and the Academic and Clinical Central Office for Research and Development (AMREC). Clinical data were collected through the Scottish Motor Neurone Disease Register (SMNDR) and the Care Audit Research and Evaluation for Motor Neurone Disease (CARE-MND) platform, with ethical approval from Scotland A Research Ethics Committee (10/MRE00/78 and 15/SS/0216). All patients provided informed consent for the use of their data for research while they were alive, and clinical and cognitive data such as ECAS and ALSFRS were collected. The owners of the data granted permission to use this dataset for secondary processing.

### Summary of Updates

The manuscript has been substantially revised throughout. The Methods section has been expanded and clarified, including a more detailed description of dataset reconstruction, feature computation, preprocessing and model evaluation. The Results section has been reorganised to present the complete performance of all machine learning models for each biomarker, replacing the previous emphasis on Random Forest and ensemble models. Additional comparisons with the baseline study have been included, together with updated tables and figures. The Discussion, limitations, conclusion and future work have been rewritten to improve interpretation of the findings and to provide a clearer distinction between the current results and their methodological limitations. Numerous revisions have also been made to improve clarity, consistency and reproducibility throughout the manuscript.

## References

1. Alzahrani, A. K., Alsheikhy, A. A., Shawly, T., Azzahrani, A. S. & AbuEid, A. I. Amyotrophic lateral sclerosis prediction framework using a multi-level encoders-decoders-based ensemble architecture technology. J. King Saud Univ. Inf. Sci. 36, 101960 (2024).

2. Smeyers, J., Banchi, E. G. & Latouche, M. C9orf72: what it is, what it does, and why it matters. Front. Cell. Neurosci. 15, 661447 (2021).

3. Pancotti, C. et al. Deep learning methods to predict amyotrophic lateral sclerosis disease progression. Sci. Reports 12, 13738 (2022).

4. Dorst, J. & Ludolph, A. C. Non-invasive ventilation in amyotrophic lateral sclerosis. Ther. Adv. Neurol. Disord. 12, 1756286419857040 (2019).

5. Louwerse, E. S., Visser, C. E., Bossuyt, P. M. M. & Weverling, G. J. The netherlands ALS consortium. Amyotrophic lateral sclerosis: mortality risk during the course of the disease and prognostic factors. J. Neurol. Sci. 152, 10–17 (1997).

6. Tafuri, B. et al. Machine learning-based radiomics for amyotrophic lateral sclerosis diagnosis. Expert. Syst. with Appl. 240, 122585 (2024).

7. Schischlevskij, P. et al. Informal caregiving in amyotrophic lateral sclerosis (ALS): a high caregiver burden and drastic consequences on caregivers’ lives. Brain Sci. 11, 748 (2021).

8. Achtert, K. & Kerkemeyer, L. The economic burden of amyotrophic lateral sclerosis: a systematic review. The Eur. J. Heal. Econ. 22, 1151–1166 (2021).

9. Feldman, E. L. et al. Amyotrophic lateral sclerosis. The Lancet 400, 1363–1380, DOI: 10.1016/S0140-6736(22)01272-7 (2022).

10. Wang, H., Guan, L. & Deng, M. Recent progress of the genetics of amyotrophic lateral sclerosis and challenges of gene therapy. Front. Neurosci. 17, 1170996, DOI: 10.3389/fnins.2023.1170996 (2023).

11. Rifai, O. M. et al. Random forest modelling demonstrates microglial and protein misfolding features to be key phenotypic markers in C9orf72-ALS. The J. Pathol. 258, 366–381 (2022).

12. Strong, M. J. et al. Amyotrophic lateral sclerosis-frontotemporal spectrum disorder (ALS-FTSD): Revised diagnostic criteria. Amyotroph. Lateral Scler. Frontotemporal Degener. 18, 153–174 (2017).

13. Abrahams, S., Newton, J., Niven, E., Foley, J. & Bak, T. H. Screening for cognition and behaviour changes in ALS. Amyotroph. Lateral Scler. Frontotemporal Degener. 15, 9–14 (2014).

14. Ileva, H., Vullaganti, M. & Kwan, J. Advances in molecular pathology, diagnosis, and treatment of amyotrophic lateral sclerosis. BMJ 383 (2023).

15. Grollemund, V. et al. Machine learning in amyotrophic lateral sclerosis: achievements, pitfalls, and future directions. Front. Neurosci. 13, 135 (2019).

16. Partners.org. PRO-ACT - home (2025). Available at: https://ncri1.partners.org/ProACT.

17. Baxi, E. G. et al. Answer als, a large-scale resource for sporadic and familial als combining clinical and multi-omics data from induced pluripotent cell lines. *Nat*. neuroscience 25, 226–237 (2022).

18. Project mine: study design and pilot analyses of a large-scale whole-genome sequencing study in amyotrophic lateral sclerosis. Eur. J. Hum. Genet. 26, 1537–1546 (2018).

19. Mazumder, S., Kiernan, M. C., Halliday, G. M., Timmins, H. C. & Mahoney, C. J. The contribution of brain banks to knowledge discovery in amyotrophic lateral sclerosis: a systematic review. Neuropathol. applied neurobiology 48, e12845 (2022).

20. Langerova, T. et al. Protein misfolding in the gastrointestinal tract predicts and prognosticates neurodegenerative disease years before symptom onset. medRxiv 2025–10, DOI: 10.1101/2025.10.00.000000 (2025). Preprint.

21. Bankhead, P. et al. Qupath: Open source software for digital pathology image analysis. Sci. reports 7, 16878 (2017).

22. Jayaraman, R. & Dutta, R. Statistical normalization methods and their impact on the classification of hyperspectral data. Pattern Recognition Letters (2019). Vol. 125, pp. 140–146.

23. Stone, M. Cross-validatory choice and assessment of statistical predictions. Journal of the Royal Statistical Society: Series B (1974). Vol. 36, No. 2, pp. 111–147.

24. Cervantes, J., Garcia-Lamont, F., Rodríguez-Mazahua, L. & Lopez, A. A comprehensive survey on support vector machine classification: Applications, challenges and trends. Neurocomputing 408, 189–215, DOI: 10.1016/j.neucom.2020.05.099 (2020).

25. Hosmer, D. W., Lemeshow, S. & Sturdivant, R. X. Applied logistic regression. Wiley (2013). 3rd edition.

26. Ng, A. Y. Feature selection, l1 vs. l2 regularization, and rotational invariance. Proceedings of the Twenty-first International Conference on Machine Learning (ICML) (2004).

27. Chen, T. & Guestrin, C. XGBoost: a scalable tree boosting system. In Proceedings of the 22nd ACM SIGKDD International Conference on Knowledge Discovery and Data Mining, 785–794, DOI: 10.1145/2939672.2939785 (ACM, San Francisco, CA, USA, 2016).

28. Du, Q., Wang, D. & Zhang, Y. The role of artificial intelligence in disease prediction: using ensemble model to predict disease mellitus. *Front*. Medicine 11, 1425305 (2024).

